# Identification of distinct trajectories in preclinical type 2 diabetes and their associations with outcomes

**DOI:** 10.1101/2024.05.07.24306979

**Authors:** Fan Yi, Jing Yuan, Fei Han, Judith Somekh, Mor Peleg, Fei Wu, Zhilong Jia, Yi-Cheng Zhu, Zhengxing Huang

## Abstract

Type 2 Diabetes Mellitus (T2DM) is increasingly prevalent and significantly impacts patients’ lives. However, the phenotypic and genetic heterogeneity of the preclinical stage of T2DM, along with the subsequent effects on various clinical outcomes, remain unclear, impeding progress in disease screening and prevention. To address this gap, we employed a robust machine learning algorithm (Subtype and Stage Inference, SuStaIn) with cross-sectional clinical data from the UK Biobank (20,305 preclinical-T2DM participants and 20,305 controls) to identify underlying subtypes and their progression trajectories for preclinical-T2DM. Our analysis revealed one subtype distinguished by elevated circulating leptin levels and decreased leptin receptor levels, coupled with increased BMI, diminished lipid metabolism, and heightened susceptibility to psychiatric conditions such as anxiety disorder, depression disorder, and bipolar disorder. Conversely, individuals in the second subtype manifested typical abnormalities in glucose metabolism, including rising glucose and HbA1c levels, with observed correlations with neurodegenerative disorders. Over ten-year follow-up observations of these individuals reveal differential deterioration in brain and heart organs, and statistically significant difference in disease risk and clinical outcomes between the two subtypes. Our findings indicate a heterogenous pathobiological basis underlying the progression of preclinical-T2DM, with clinical implications for understanding human health from a multiorgan perspective, and improving disease risk screening, prediction, and prevention efforts.

## Introduction

Type 2 Diabetes Mellitus (T2DM) represents a significant global public health challenge, with its prevalence steadily increasing. In 2021, it affected 537 million adults worldwide ^1^. Projections from the International Diabetes Federation indicate that by 2030, this number will rise to 643 million, reaching a staggering 783 million by 2045 ^1,2^. The phase that clinical symptoms remain absent but biological irregularities hint at the potential development of T2DM, called as preclinical stage of Type 2 Diabetes (hereafter, preclinical-T2DM), play an important role in the developing into T2DM ^3^. Large cohort studies, such as the UK Biobank (UKB), offer a valuable opportunity to investigate this pivotal phase. Within these cohorts, individuals are identified as being at risk of T2DM development and are monitored for progression during follow-up. Understanding the heterogeneity of preclinical-Type 2 Diabetes will be beneficial to the precision prevention and early diagnosis of heterogeneous T2DM.

The heterogeneity and organ-specific impacts of preclinical-T2DM remain poorly understood, though these related with T2DM have been widely recognized ^4,5^. First, there is less knowledge regarding how preclinical-T2DM progresses over time before the onset of T2DM. Second, it remains unclear how the phenotypic and genetic etiology of preclinical-T2DM vary. Third, the magnitude and extent of interindividual differences in progression trajectories of preclinical-T2DM and their alterations on clinical outcomes, such as cardiovascular, renal, ophthalmological, neurological, and cognitive functions, remain undetermined. Existing studies have clustered prediabetes into subtypes regardless the progressive stage of prediabetes and explored their associations with T2DM as well as its complications ^6^, but such approaches fail to capture the longitudinal heterogeneity of preclinical-T2DM. Conversely, the progressive heterogeneity is a hallmark of T2DM, which could be sourced from the preclinical stage of the T2DM continuum.

Addressing this challenge can be achieved by leveraging machine learning models that are increasingly used in biomedical research ^7^. One such model, the subtype and stage inference (SuStaIn) model ^8^, originally designed to capture disease progression patterns in chronic conditions, facilitates longitudinal inference from cross-sectional data by automatically identifying distinct spatiotemporal trajectories of cumulative pathological alterations shown by measured biomarkers ^8–13^. In this study, we employed SuStaIn to decipher heterogenous progressive patterns of preclinical-T2DM, offering valuable insights into disease onset and progression. This aids in the establishment of quantitative metrics for T2DM screening and prognostication. By identifying the disease pathways and subtypes of preclinical-T2DM, and exploring systematic changes in the brain, heart, and other clinical outcomes linked to preclinical-T2DM, we can enhance our ability to precisely assess it in clinical practice. This not only benefits individuals with preclinical-T2DM but also contributes to better health outcomes, reducing the risk of neuropathy, cognitive dysfunction, and cardiovascular issues, etc ^14^.

In the present study, we embarked on a comprehensive investigation into the heterogenous progression of preclinical-T2DM and its implications for clinical outcomes using a multi-faceted research approach (Fig. 1). First, we identified 20,305 preclinical-T2DM subjects with a balanced 20,305 control group from UKB for analysis (Fig. 1a). Next, we utilized screened 18 preclinical-T2DM-associated clinical indexes and applied SuStaIn to stratify preclinical-T2DM subjects into distinct subtypes and stages, leading to two subtypes with distinct progression trajectories (Fig. 1b). Subsequently, we analysed phenotypic associations between the two subtypes and a variety of clinical outcomes, including cardiac diseases, kidney diseases, brain disorders, and cognitive functions as well as molecular phenotypes, proteins and metabolites (Fig. 1c). Furthermore, we identified significantly subtype-specific genetic variants associated with the two preclinical-T2DM subtypes utilizing Genome-Wide Association Studies (GWAS). Expending on the findings from GWAS, we investigated the genetic relationships between the two subtypes and clinical outcomes via genetic correlation analysis, genetic colocalization and Mendelian Randomization (MR) analysis (Fig. 1c). By exploring the genetic and molecular landscape of preclinical-T2DM subtypes and their impact on clinical outcomes, we highlighted how underlying phenotypic and genetic variation drives the subtypes and stages of preclinical-T2DM. These insights improve our understandings of the complex interplay between preclinical-T2DM-associated metabolic disorders and human health. Our findings pave the way for personalized medicine approaches in the prevention and management of T2DM and its effects on subsequent clinical outcomes.

**Fig. 1:**
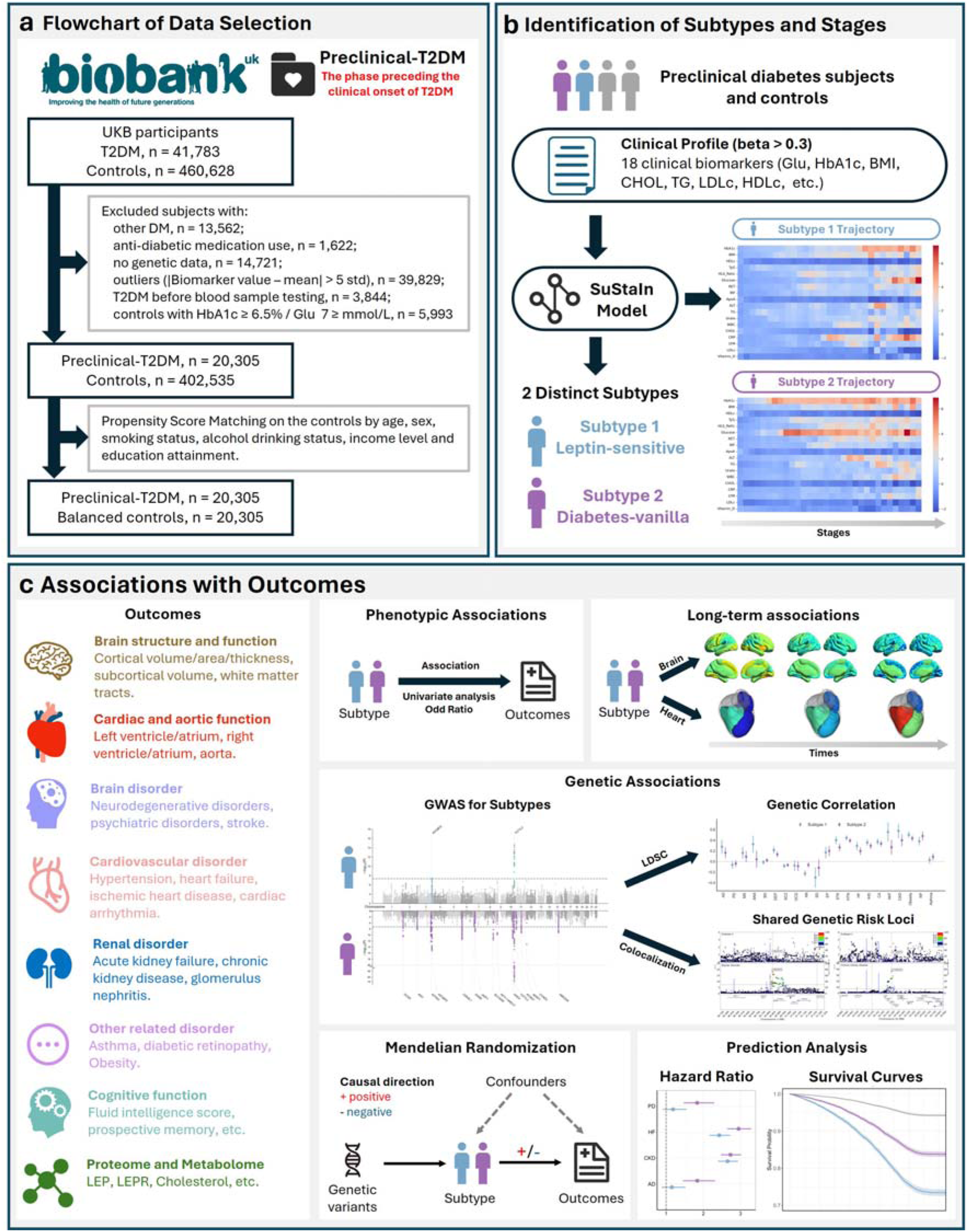
Overview of the study design. **a**. Flow chart depicting the inclusion and exclusion criteria for data selection from the UK Biobank (UKB). We selected subjects with preclinical-T2DM, defined as the phase that preceding the onset of T2DM, along with a corresponding balanced control group from the UKB for analysis. **b.** Application of the SuStaIn model for identification of subtypes and stages for preclinical-T2DM. The model identified two subtypes, Subtype 1 (Leptin-sensitive), and Subtype 2 (Diabetes-vanilla) with two distinct progression trajectories, using preclinical-T2DM-associated 18 clinical biomarkers. **c.** Associations with outcomes. We examined the associations of the identified subtypes with a variety of outcomes using multiple analytical methods, including phenotypic associations, long-term imaging-derived phenotypes association analysis, genetic associations, Mendelian randomization, and prediction analysis.

## Results

### Identification of robust subtype and stages for preclinical-T2DM

Two distinct subtypes of preclinical-T2DM were identified with 18 biomarkers of 40,610 individuals in UKB using the SuStaIn algorithm (Fig. 2a). We selected preclinical-T2DM individuals, defined at the individuals at the cohort baseline who will have T2DM in the follow-up based on the inclusion and exclusion criteria (Fig. 1a), resulting in 20,305 preclinical-T2DM and 20,305 propensity score matched controls. Due to the computation requirements, we selected 18 out of 62 biomarkers from the baseline UKB dataset due to their significance for preclinical-T2DM, as determined by larger effect sizes in univariable logistic regression (Fig. 2a and Table 1). Using a ten-fold cross-validation approach, we determined the most robust result, revealing two distinct subtypes, Subtype 1 (S1) and Subtype 2 (S2), with 36 subtype-specific stages (S1, N= 9,402, mean age 59 years, 52% female, and S2, N = 10,903, mean age 60 years, 37% female). This distinction highlights two distinct clinical and pathophysiological trajectories for preclinical-T2DM (Fig. 2a-b). Moreover, cross-validation demonstrated a high consistency in the identification of subtypes for each preclinical-T2DM participants, with the majority of subjects (96.18% on average) consistently assigned to the same group across validation folds (Supplementary Fig. 2). These findings further confirm the stability and reproductivity of the results obtained from SuStaIn.

**Fig. 2:**
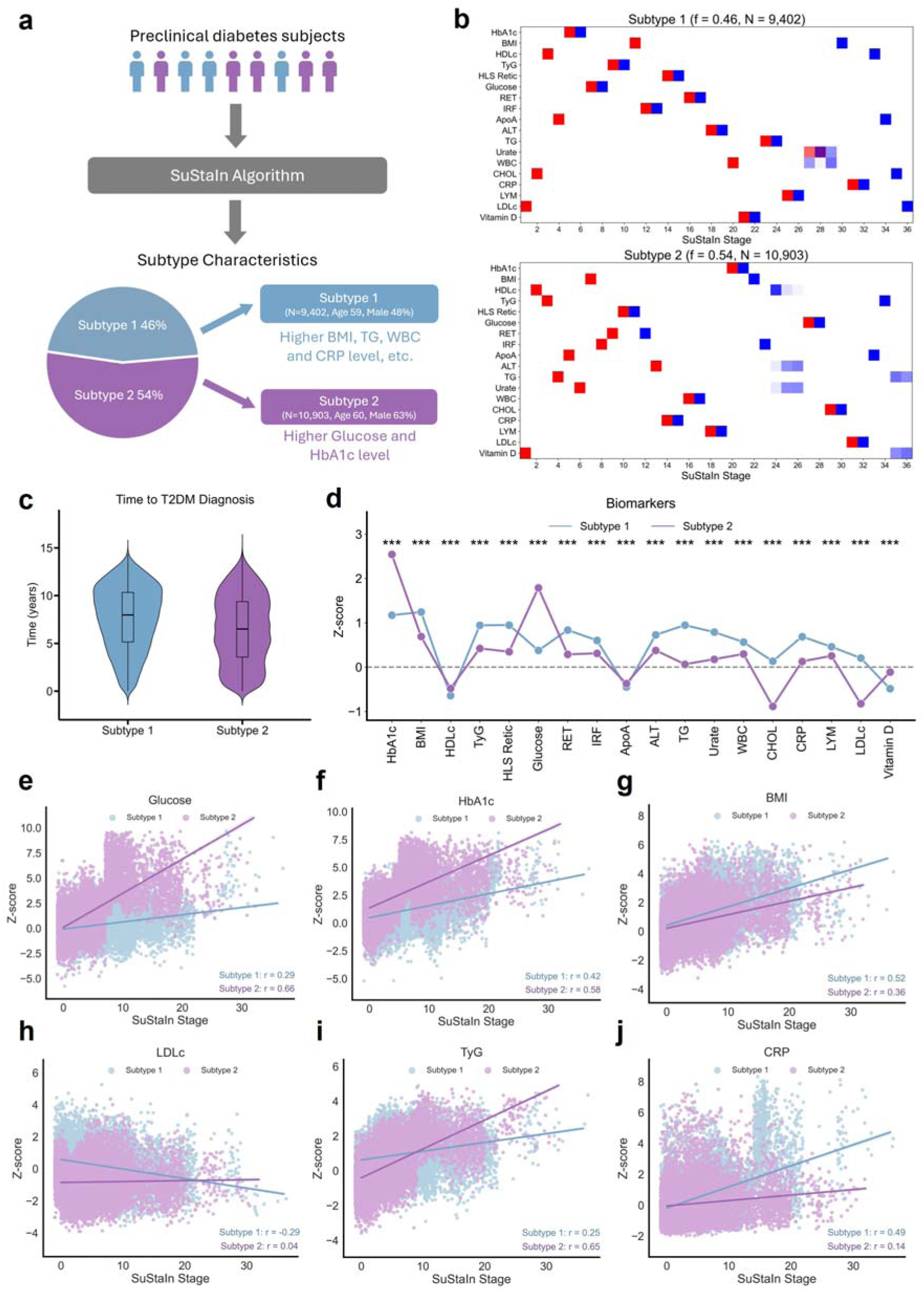
Identification of subtypes and stages for preclinical-T2DM. **a.** An overview of the two distinct preclinical-T2DM subtypes identified by the SuStaIn algorithm. **b.** Positional variance diagrams of two distinct metabolic trajectories obtained from SuStaIn. The diagrams visualize the cumulative probability of each biomarker reaching a specific z-score, depicted in different colours. Red indicates mild effects (z-score = 1, i.e., 1 standard deviation from the healthy control average) and blue indicates severe effects (z-score = 2). The colour density represents the proportion of the posterior distribution where events (y-axis) occur at specific positions in the sequence (x-axis); f represents the proportion of individuals assigned to each phenotype. **c.** Comparison of time to T2DM diagnosis for the two subtypes. **d.** Comparison of the mean z-scores of 18 selected clinical biomarkers across the two identified subtypes, where biomarkers were z-scored relative to the control group, adjusting for age, sex, smoking status, alcohol drinking status, income level, and educational attainment. A higher z-score indicates a greater deviation from the control group norm. **e-j**. Progressions of various biomarkers across SuStaIn stages. We illustrated 6 selected biomarkers with remarkably distinct progression on the two subtypes. Progressions of other biomarkers were presented in Supplementary Figs. 5-16. R is the Pearson’s correlation between biomarkers and SuStaIn stages for each subtype.

**Table 1.**
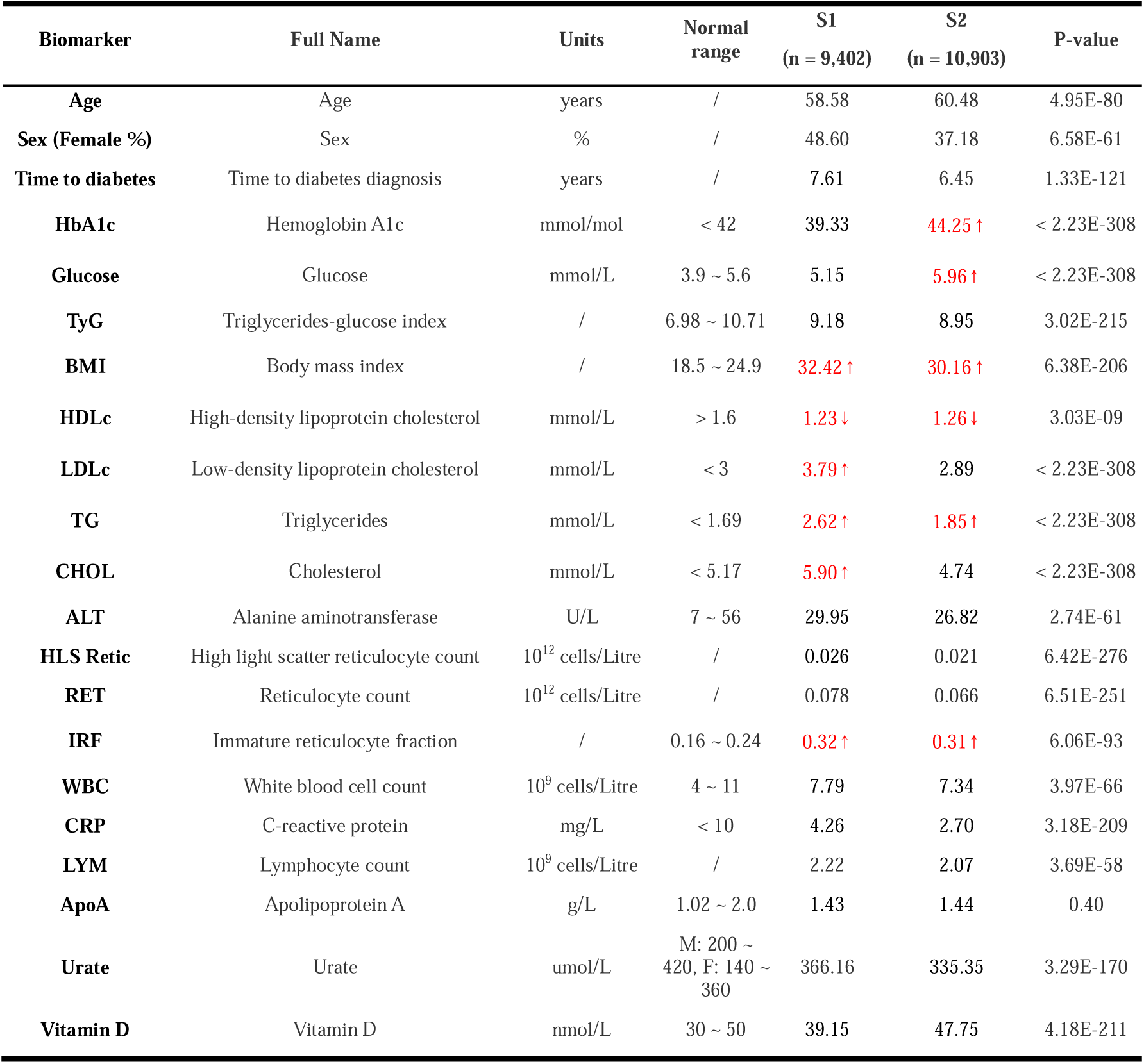
Basic characteristics and 18 clinical biomarkers of the two subtypes of preclinical-T2DM. Abnormal values of clinical indexes are marked in red colour.

| Biomarker | Full Name | Units | Normal range | S1<br>(n = 9,402) | S2<br>(n = 10,903) | P-value |
| --- | --- | --- | --- | --- | --- | --- |
| Age | Age | years | / | 58.58 | 60.48 | 4.95E-80 |
| Sex (Female %) | Sex | % | / | 48.60 | 37.18 | 6.58E-61 |
| Time to diabetes | Time to diabetes diagnosis | years | / | 7.61 | 6.45 | 1.33E-121 |
| HbA1c | Hemoglobin A1c | mmol/mol | < 42 | 39.33 | 44.25 ↑ | < 2.23E-308 |
| Glucose | Glucose | mmol/L | 3.9 ~ 5.6 | 5.15 | 5.96 ↑ | < 2.23E-308 |
| TyG | Triglycerides-glucose index | / | 6.98 ~ 10.71 | 9.18 | 8.95 | 3.02E-215 |
| BMI | Body mass index | / | 18.5 ~ 24.9 | 32.42 ↑ | 30.16 ↑ | 6.38E-206 |
| HDLc | High-density lipoprotein cholesterol | mmol/L | > 1.6 | 1.23 ↓ | 1.26 ↓ | 3.03E-09 |
| LDLc | Low-density lipoprotein cholesterol | mmol/L | < 3 | 3.79 ↑ | 2.89 | < 2.23E-308 |
| TG | Triglycerides | mmol/L | < 1.69 | 2.62 ↑ | 1.85 ↑ | < 2.23E-308 |
| CHOL | Cholesterol | mmol/L | < 5.17 | 5.90 ↑ | 4.74 | < 2.23E-308 |
| ALT | Alanine aminotransferase | U/L | 7 ~ 56 | 29.95 | 26.82 | 2.74E-61 |
| HLS Retic | High light scatter reticulocyte count | 10<sup>12</sup> cells/Litre | / | 0.026 | 0.021 | 6.42E-276 |
| RET | Reticulocyte count | 10<sup>12</sup> cells/Litre | / | 0.078 | 0.066 | 6.51E-251 |
| IRF | Immature reticulocyte fraction | / | 0.16 ~ 0.24 | 0.32 ↑ | 0.31 ↑ | 6.06E-93 |
| WBC | White blood cell count | 10<sup>9</sup> cells/Litre | 4 ~ 11 | 7.79 | 7.34 | 3.97E-66 |
| CRP | C-reactive protein | mg/L | < 10 | 4.26 | 2.70 | 3.18E-209 |
| LYM | Lymphocyte count | 10<sup>9</sup> cells/Litre | / | 2.22 | 2.07 | 3.69E-58 |
| ApoA | Apolipoprotein A | g/L | 1.02 ~ 2.0 | 1.43 | 1.44 | 0.40 |
| Urate | Urate | umol/L | M: 200 ~ 420, F: 140 ~ 360 | 366.16 | 335.35 | 3.29E-170 |
| Vitamin D | Vitamin D | nmol/L | 30 ~ 50 | 39.15 | 47.75 | 4.18E-211 |

### Two distinct metabolic trajectories of preclinical-T2DM

The two subtypes of preclinical-T2DM exhibited significant differences in clinical biomarkers (Fig. 2d). Compared to S2, S1 exhibited elevated levels of Body Mass Index (BMI), total cholesterol (CHOL), urate, and low-density lipoprotein cholesterol (LDLc) (Table 1, Figs. 2c, g, h, and Supplementary Figs 12, 14). These heightened biomarkers may suggest a correlation with more severe metabolic dysregulation in lipid metabolism for S1. Additionally, S1 demonstrated higher levels in inflammatory biomarkers, including reticulocyte count (RET), immature reticulocyte fraction (IRF), C-reactive protein (CRP), and lymphocytes (LYM) (Figs. 2c, j, and Supplementary Figs 7-8, 13, 15), potentially indicating a more pronounced inflammatory activity contributing to the progression of S1. In contrast, S2 exhibited elevated levels of glucose as well as HbA1c (Fig. 2c). Notably, the rates of progression for glucose and HbA1c were significantly faster in S2 compared to S1 (Fig. 2d, e-f), suggesting a more rapid decline in glycaemic control. Moreover, S1 presented with lower levels of Vitamin D (Fig. 2c and Table 1). Studies have reported that vitamin D supplementation among individuals with prediabetes can mitigate the risk of developing T2DM and facilitate the transition from prediabetes to normoglycemia ^15^. Furthermore, it was observed that S2 exhibited a more rapid progression to T2DM through stages, with an average progression time of 6.5 years, compared to the 7.6 years observed for S1 (Fig. 2c, Table 1, and Supplementary Fig. 4). This observation suggests that although both subtypes were in the early stage of T2DM, S2 may represent a more aggressive progression to T2DM, possibly due to faster deterioration of glycaemic control. These findings underscore the heterogeneity within preclinical-T2DM and suggest that the two subtypes may represent distinct pathological progressions of T2DM.

### Phenotypic associations of preclinical-T2DM subtypes on outcomes

We examined the phenotypic association between the two subtypes and 22 types of diseases, grouping into 12 brain, 4 cardiovascular, 3 renal, and 3 other related disorders (Fig. 3a and Supplementary Table 3), to explore clinical significance of the two preclinical-T2DM subtypes. We discovered both subtypes of preclinical-T2DM are associated with almost all the cardiovascular diseases, renal diseases, asthma, obesity, and diabetic retinopathy (Fig. 3a). Notably, S1 was associated with a significantly higher risk of obesity compared to S2 (OR_s1_ = 6.02, p_s1_ < 2.23E-308, and OR_s2_ = 3.02, p_s2_ = 6.39E-219). We also observed significant disparities in brain disorders between two subtypes. Compared with S2, S1 showed a significantly higher risk of psychiatric disorders, including anxiety disorder (OR_s1_ = 2.17, p_s1_ = 5.25E-72, and OR_s2_ = 1.49, and p_s2_ = 2.58E-18), bipolar disorder (OR_s1_ = 3.33, p_s1_ = 7.21E-16, and OR_s2_= 1.98, p_s2_ = 5.99E-05), depression disorder (OR_s1_ = 2.33, p_s1_ = 1.14E-128, and OR_s2_ = 1.55, p_s2_ = 1.65E-32), obsessive compulsive disorder (OR_s1_ = 2.57, p_s1_= 0.0069, OR_s2_ = 1.40, p_s2_ = 0.0080) and sleep disorder (OR_s1_= 4.29, p_s1_ = 2.01E-153, and OR_s2_ = 2.86, p_s2_ = 2.81E-70). S2, in contrast, was more strongly associated with neurodegenerative disorders, including Alzheimer’s disease (OR_s1_= 1.06, p_s1_ = 0.65, and ORs_2_ = 1.51, p_s2_ = 1.11E-04), Parkinson’s disease (OR_s1_= 0.92, p_s1_ = 0.53, and OR_s2_ = 1.48, p_s2_ = 4.30E-05), and a more increased risk of stroke (OR_s1_= 1.62, p_s1_ = 5.52E-10, and OR_s2_ = 2.08, p_s2_ = 3.37E-26). These associations highlight the intricate relationship between preclinical-T2DM subtypes and a wide array of health conditions, shedding light on potential shared mechanism-related pathways and comorbidities.

**Fig. 3:**
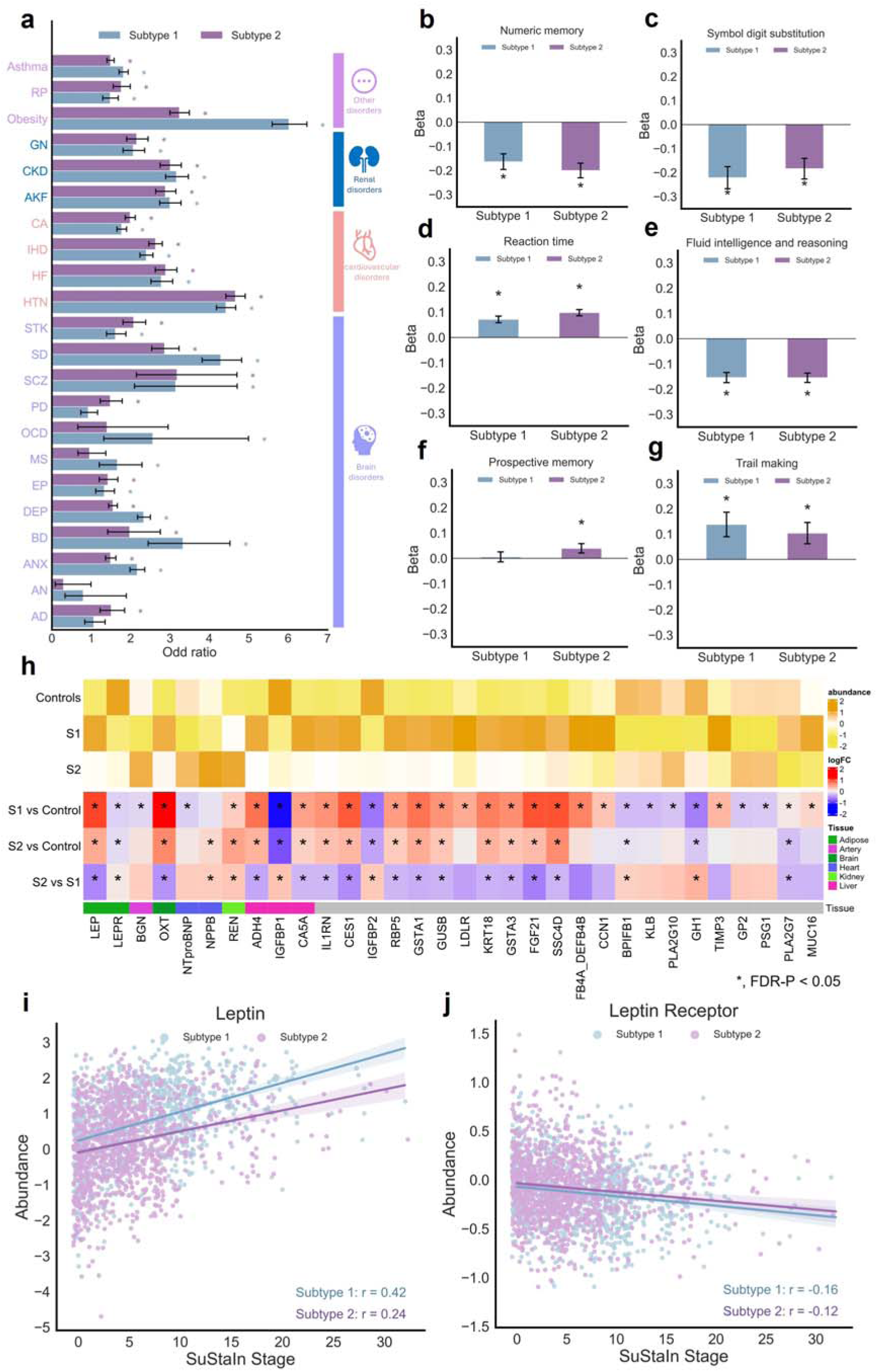
Phenotypic associations with outcomes. **a.** The odds ratios (ORs) of associations between preclinical-T2DM subtypes and 22 diseases, including 12 brain disorders, 4 cardiac disorders, 3 renal disorders, and 3 other related disorders. The significance of each association was calculated using a t-test, with “*” indicating significant differences after adjusting for multi-testing by Benjamini-Hochberg procedure to control the false discovery rate (FDR) at the 5% level (FDR-p < 0.05). **b-g.** Phenotypic associations between subtypes and 6 cognitive functions, which were z-scored relative to the control group and adjusted for sex, age, smoking status, alcohol drinking status, income level, and educational attainment. Associations were measured using univariate analysis, where “*” indicates a significant association after FDR correction (FDR-p < 0.05). **h.** Comparison of proteomic expressions between controls and the two subtypes. Proteins from six tissues: adipose, artery, brain, heart, kidney, and liver, were analysed. Log fold change (LogFC) was used to identify differences in proteomic expressions across the three groups: Controls, S1 and S2. Significant differences were assessed using one-way ANOVA tests with FDR-p < 0.05. **I-j.** Progressions of the abundance of leptin (LEP) and leptin receptor (LEPR) across SuStaIn stages. Abbreviations for diseases: AD, Alzheimer’s disease; AN, anorexia nervosa; ANX, anxiety disorder; BD, bipolar disorder; DEP, depression disorder; EP, epilepsy; MS, multiple sclerosis; OCD, obsessive compulsive disorder; PD, Parkinson’s disease; SCZ, schizophrenia; SD, sleep disorder; STK, stroke; HTN, hypertension; HF, heart failure; IHD, ischemic heart disease; CA, cardiac arrhythmias; AKF, acute kidney failure; CKD, chronic kidney disease; GN, glomerulus nephritis; RP, diabetic retinopathy.

We also investigated the associations between two subtypes and 6 cognitive functions tested in the UKB. Both subtypes were associated with declines across several cognitive aspects, including numeric memory, symbol digit substitution, reaction time, fluid intelligence and reasoning, and trail making test performance (Figs. 3b-g). Specifically, S1 showed a more pronounced effect on executive functioning, with greater impairments in symbol digit substitution (Fig. 3c, β_s1_ = −0.22, p_s1_ = 4.84E-07, and β_s2_ = −0.18, p_s2_ = 2.07E-05) and Trail making tests (Fig. 3g, β_s1_= 0.14, p_s1_ = 0.0044, and β_s2_ = 0.10, p_s2_ = 0.014). Conversely, S2 showed a more substantial impact on numeric memory (Fig. 3b, β_s1_= −0.16, p_s1_ = 4.84E-07, and β_s2_ = −0.20, p_s2_ = 6.63E-11) and reaction time (Fig. 3d, β_s1_= 0.07, p_s1_ = 3.24E-08, and β_s2_ = 0.10, p_s2_ = 1.93E-15), involving the ability of memorising and neural processing speed and response.

We evaluated the proteomic phenotypes with two subtypes of preclinical-T2DM. In the proteomic analysis, we found heart-specific and kidney-specific proteins (NTproBNP, NPPB and REN) were highly expressed in S2, while brain-specific protein, OXT, are highly expressed in S1 (Fig. 3h). Fibroblast Growth Factor 21 (FGF21) is upregulated in both subtypes, with the highest expression observed in S1 compared to healthy controls. FGF21, a pivotal player in the regulation of energy balance and glucose as well as lipid homeostasis, has gathered attention as a therapeutic target for T2DM and obesity. Clinical trials utilizing FGF21 analogues and mimetics have shown promise in patients with obesity and T2DM ^16^, suggesting a potential physiological response of FGF21 to the preclinical status of T2DM. Leptin (LEP) exhibits a similar expression pattern to FGF21, indicating a potential association with the preclinical state of T2DM (Figs. 3h-i). Conversely, the expression patterns of Leptin Receptor (LEPR) across the three groups are dramatically opposite, suggesting a possible impairment in leptin receptor signalling (Figs. 3h-j). This observation aligns with studies utilizing leptin receptor-deficient db/db mice as models for T2DM ^17^. Elevated circulating leptin concentrations, as observed in individuals with obesity, are often attributed to leptin resistance ^18,19^, potentially implicating S1 is related with leptin resistance. Leptin-resistant syndromes are known to contribute to severe insulin resistance and diabetes ^20^. Furthermore, Insulin-like Growth Factor Binding Proteins 1 and 2 (IGFBP1 and IGFBP2) as well as β-klotho (KLB) exhibit decreased expression levels in S1 compared to healthy controls (Fig. 3h). Reduced concentrations of circulating IGFBP1 and IGFBP2 have been linked to insulin resistance and diabetes ^21^, while KLB, acting as a cell-surface glucose sensor and co-receptor for FGF21, holds promise as a therapeutic target for T2DM by modulating glucose-stimulated insulin release in pancreatic β-cells. As the ratio of triglycerides to high-density lipoprotein cholesterol (TG: HDLc) and the metabolic score for insulin resistance (METS-IR) serve as readily measurable markers of insulin resistance, our findings reveal that S1 exhibits significantly elevated TG: HDLc and METS-IR levels compared to S2 and the healthy control group. Significantly higher *IL1RN* is observed on S1, compared with S2 and controls. Circulating IL-1RA (encoded by *IL1RN*), an endogenous inhibitor of proinflammatory IL-1β, may be protective against the development of insulin resistance ^22^.

Moreover, we evaluated the associations between metabolomic phenotypes and two subtypes of preclinical-T2DM. We observed significantly higher levels of lipoproteins, such as large VLDL, very large VLDL, chylomicrons, and extremely large VLDL, in S1 compared to S2, and the healthy control group (Supplementary Fig. 17). Previous studies have also reported positive associations of T2DM with lipoprotein subfractions in large VLDL, very large VLDL, and chylomicrons and extremely large VLDL ^23^. Our findings suggested that these associations tended to be particularly pronounced in S1. Furthermore, we found that levels of large HDL, fatty acid ratios, and chylomicrons and extremely large VLDL ratios are lowest in S1 among the three groups. This indicates an elevated risk of cardiovascular disease for S1 individuals with preclinical-T2DM.

### Long-term effects of preclinical-T2DM subtypes on the brain and the heart

The analysis of long-term effects on brain and heart structure and function across the two subtypes unveiled distinct outcomes in both organs (Fig. 4). Subtypes were grouped into three categories based on the intervals of imaging acquisition following their initial assessment at the UK Biobank: 4-7 years, 7-10 years, and more than 10 years, ensuring a balanced sample size per group. In terms of MRI-based brain structure, S1 exhibited significant atrophy in volumes of brain regions and cortical thickness over time, particularly in the left hippocampus, right accumbens, left thalamus, and putamen (Figs. 4a, c, and Supplementary Figs. 18, 20). Conversely, S2 displayed notable atrophy in brain region volumes and areas of the right paracentral, left fusiform, and superior frontal regions (Figs. 4a, b, and Supplementary Figs. 18-19). Regarding white matter integrity and microstructural organization, both subtypes showed significant decreases in mean white matter fractional anisotropy (FA) in regions such as the middle cerebellar peduncle, cerebral peduncle, and fornix regions (Figs. 4d and Supplementary Fig 21). S1 also demonstrated marked reductions in the pontine crossing tract and right external capsule, while S2 displayed significant decreases in the splenium of the corpus callosum and the right retrolenticular part of the internal capsule (Fig. 4d).

**Fig. 4:**
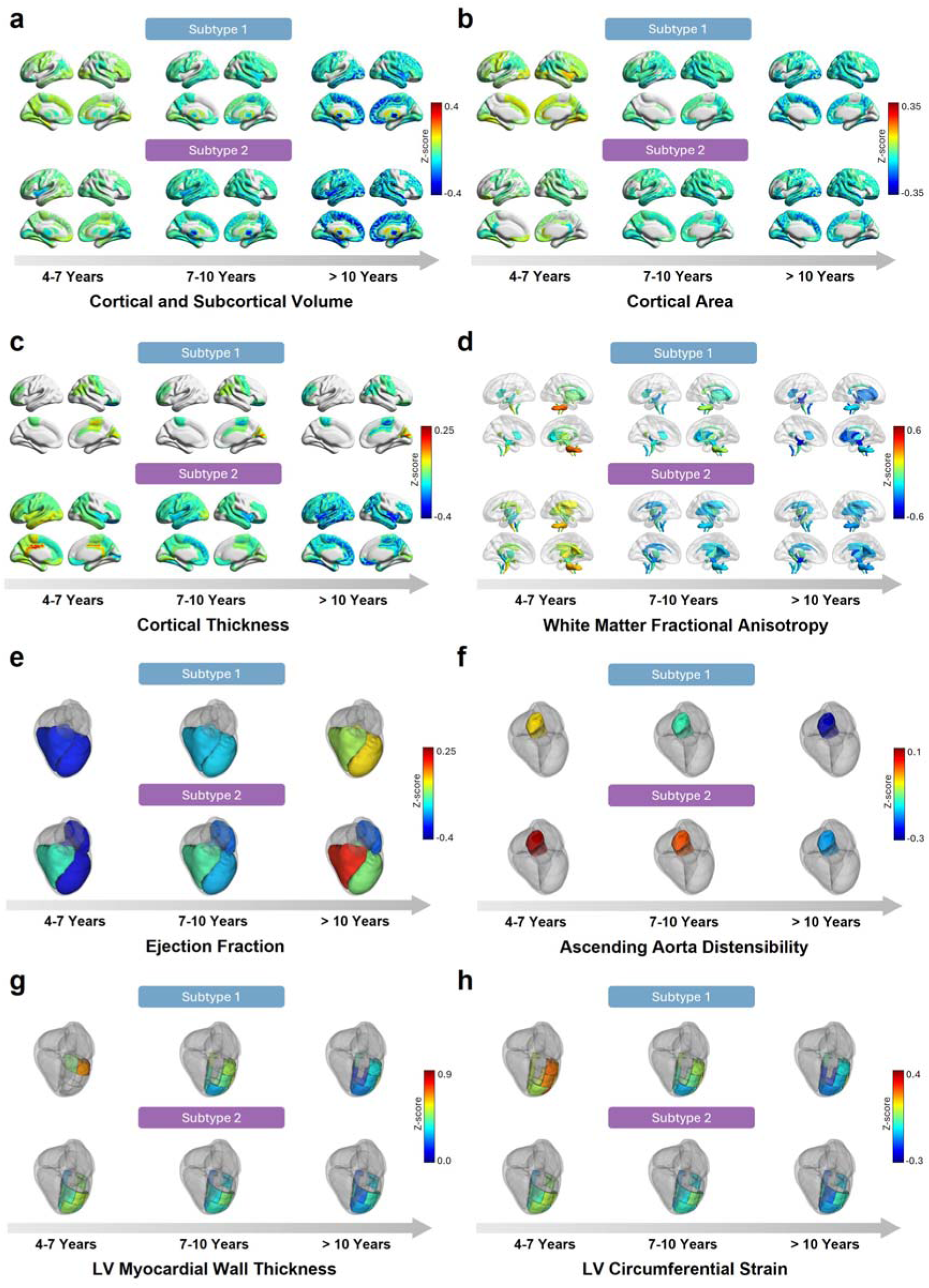
Long-term associations with the brain and the heart. **a-d.** Long-term effects of preclinical-T2DM subtypes on the selected image derived phenotypes (IDPs) from brain MRIs. **e-h**. Long-term effects of preclinical-T2DM subtypes on the selected IDPs from CMRs. Subjects for each subtype were categorized into three groups based on imaging acquisition intervals: 4-7 years, 7-10 years, and more than 10 years, to analyse temporal changes in brain and heart structures and functions. We illustrated selected IDPs with most 11 pronounced effect associated with subtypes. Long-term associations with other IDPs were presented in Supplementary Figs. 28-33. The IDP values were z-scored relative to the control group and adjusted for sex, age, smoking status, alcohol drinking status, income level, and educational attainment. The colormap illustrates the mean z-scores of the IDPs for each time interval, where a red colour indicates larger IDP values, and a blue colour indicates smaller IDP values.

Concerning cardiac magnetic resonance (CMR) traits, both S1 and S2 demonstrated an increase in ejection fraction in the left ventricle (LV) and right ventricle (RV), alongside a significant decrease in ascending aorta distensibility (Fig. 4e-f). Additionally, S2 exhibited a mild increase in ejection fraction in the left atrium (LA) (Fig. 4e). Moreover, S1 displayed a marked decrease in circumferential strain in the LV American Heart Association (AHA) segments 7, 9, 11, 14, and 16 (Fig. 4h), while S2 exhibited a notable reduction in myocardial wall thickness in LV AHA segments 8, 13, 14, and 15 (Fig. 4g).

### GWAS on the two preclinical-T2DM subtypes

We investigated the subtype-specific associated SNPs for the two preclinical-T2DM subtypes using GWAS, respectively, identifying 2 genomic risk loci with 2 independent lead SNPs for S1, and 15 genomic risk loci with 19 lead SNPs for S2 (Fig. 5a, and Supplementary Tables 5-6). The SNP-based heritability estimates for the two subtypes are 0.14 and 0.17, respectively (Supplementary Fig 34). Notably, the GWAS results for the subtypes are weakly correlated (Pearson correlation coefficient 0.34, p < 2.23E-308) though significantly, indicating a minimal correspondence with the presence of some shared genetic variants while also underscoring substantial differences (Supplementary Fig. 35). The gene annotation of these lead SNPs reveals that there is obvious difference between the two subtypes. A S1-assoicated significant SNP at 3q27.2, rs66513933, is in the intron of *IGF2BP2*, the high concentration of which is strongly associated with low type 2 diabetes risk ^24^. Another risk loci of S1 at 10q25.2-q25.3 with 3 independent significant SNPs, are in the intron of *TCF7L2*, the most potent locus for T2DM ^25^. Almost all the genes associated with the lead SNPs of S2 are reported associated with T2DM before, including *IGF2BP2* and *TCF7L2*. For example, *GCKR* is a hepatocyte-specific inhibitor of the glucose-metabolizing enzyme glucokinase ^26^; *IRS1* plays a critical role in insulin-signalling pathways ^27^; A paralog of *ELF5A2* is associated with *T2DM* ^28^; *CDKAL1* are involved in misfolded insulin, leading to oxidative and ER stress in the pancreatic β-cells ^29^; *DGKB* is causally associated with T2DM ^30^; *JAZF1* directly and negatively regulates insulin gene transcription ^31^; a loss-of-function of *SLC30A8* protects against T2DM ^32^; *HHEX* are repeatedly associated with T2DM ^33^; *KCNQ1* is highly associate with the risk of T2DM ^34^; ARAP1 is located near risk alleles for T2DM ^35^; a variant of *CCND2* could reduce risk of T2DM by half ^36^; *HMG20A* is a key for the functional maturity of islet β cell ^37^. The consistence between our results and these reported associations indicates the reliably of our GWASs results for the two subtypes. Meanwhile, the large difference between the GWASs of S1 and S2 showed the different genetic sources of the two subtypes, suggesting the rationality of our subtyping results with SuStaIn, which forms a foundation for the genetic association with the clinical outcomes.

**Fig. 5:**
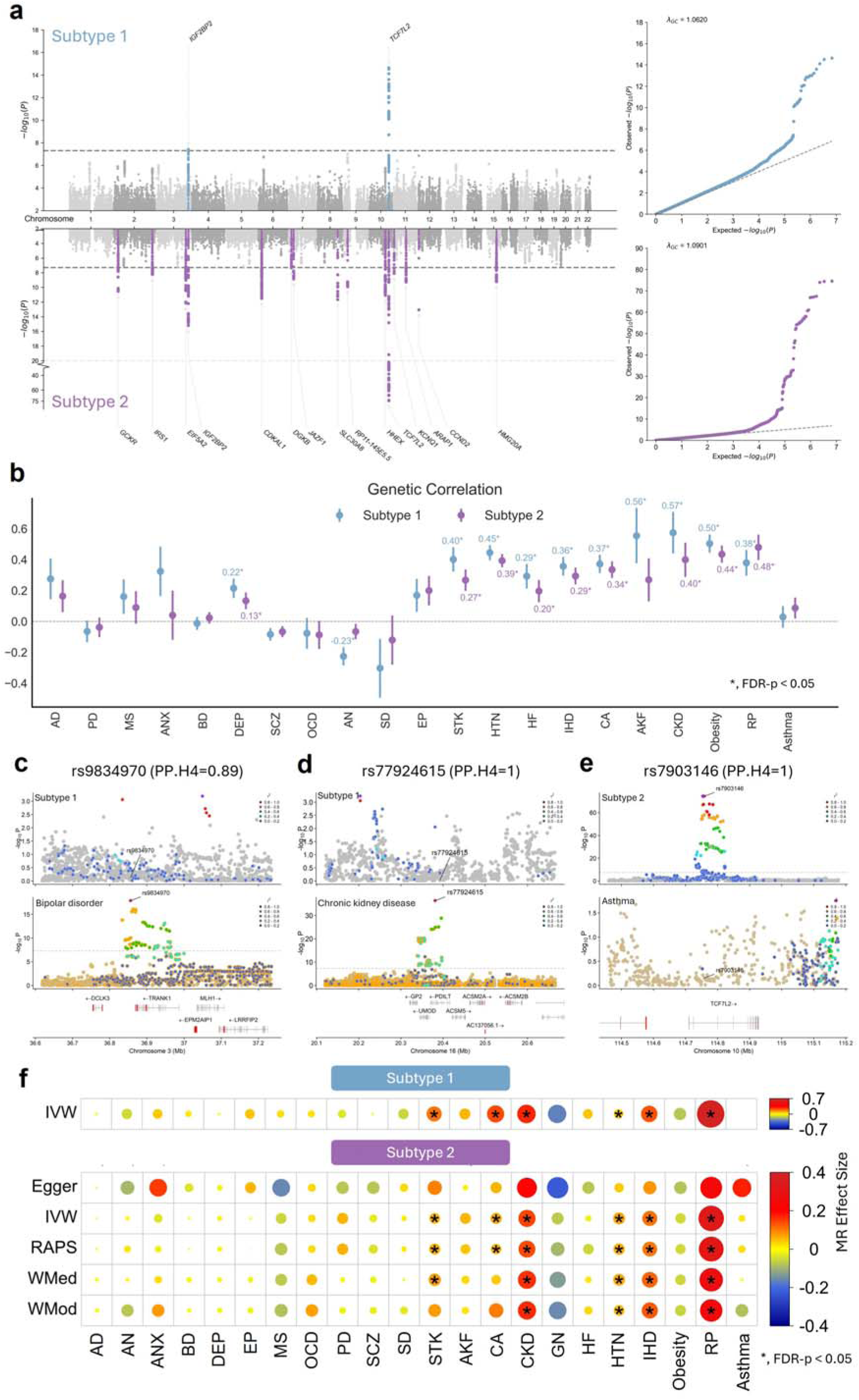
Genetic profiles and associations for the preclinical-T2DM subtypes with outcomes. **a.** The Miami and Q-Q plot of GWAS results for the two preclinical-T2DM subtypes. The dashed line indicated the significant level (p < 5E-08). Regions in a sidling window size of 500 kilobase (kb) around the lead SNPs were highlighted in the plot. Genes annotated for lead SNPs were marked in each region on the GWAS plot. The genomic control lambda (λ_GC_) on the QQ-plot is used to assess the degree of inflation in test statistics due to potential population stratification. A value of λ_GC_ close to 1 indicates no significant bias from population stratification. **b.** Genetic correlations between subtypes and 22 diseases. Results that passed the significant threshold adjusted by Benjamini-Hochberg procedure to control the FDR at the 5% level (FDR-p < 0.05) were marked in the plot. **c-e.** Significant colocalization results between subtypes and diseases (PPH4 > 0.75). **f.** Genetic causal effects estimated by MR analyses of subtypes on 22 diseases. we employed the inverse variance weighted (IVW) method for S1 with 2 SNPs as instrument variables (IV), and another four MR methods, MR egger (Egger), MR-RAPS (RAPS), weighted median (WMed) and weighted mode (WMod) for S2 with 18 SNPs as IVs. Results that pass the significant threshold adjusted by Benjamini-Hochberg procedure to control the FDR at the 5% level (FDR-p < 0.05) were marked with asterisks. Abbreviations for diseases: AD, Alzheimer’s disease; AN, anorexia nervosa; ANX, anxiety disorder; BD, bipolar disorder; DEP, depression disorder; EP, epilepsy; MS, multiple sclerosis; OCD, obsessive compulsive disorder; PD, Parkinson’s disease; SCZ, schizophrenia; SD, sleep disorder; STK, stroke; HTN, hypertension; HF, heart failure; IHD, ischemic heart disease; CA, cardiac arrhythmias; AKF, acute kidney failure; CKD, chronic kidney disease; GN, glomerulus nephritis; RP, diabetic retinopathy.

### Genetic associations between preclinical-T2DM subtypes and outcomes

We examined the genetic correlation between the two preclinical-T2DM subtypes and a series of disease outcomes using linkage disequilibrium score regression (LDSC). Both subtypes showed significant genetic associations with a range of brain disorders (for example, depression disorder and stroke), cardiac disorders (hypotension, heart failure, ischemic heart disease and cardiac arrhythmias), renal disorders (acute kidney failure and chronic kidney disease) and other disorders (obesity and diabetic retinopathy) (Fig. 5b). Notably, both subtypes exhibited strong genetic correlations with hypertension (rg_s1_ = 0.44, p_s1_ = 4.69E-20, and rg_s2_ = 0.39, p_s2_ = 1.08E-19), chronic kidney disease (rg_s1_= 0.57, p_s1_ = 1.94E-05, and rg_s2_ = 0.40, p_s2_ = 0.00030), and obesity (rg_s1_= 0.50, p_s1_ = 1.65E-17, and rg_s2_ = 0.44, p_s2_ = 2.13E-15), suggesting a shared genetic predisposition that may increase the risk of these diseases (Fig. 5b). Alzheimer’s disease, anxiety disorder, schizophrenia, anorexia nervosa and acute kidney failure are significantly associated with S1, but not with S2. Particularly, acute kidney failure was significantly association with only S1 (rg_s1_= 0.56, p_s1_ = 0.0019, and rg_s2_ = 0.27, p_s2_ = 0.052) and chronic kidney disease has slightly higher genetic correlation with S1 than S2, suggesting a distinct genetic component influencing renal outcomes (Fig. 5b). Furthermore, S2 showed a significantly genetic correlation with epilepsy compared to S1 (Fig. 5b). These subtype-specific genetic association of different diseases reveals different genetic risk on diabetes-related complications.

For cognitive traits, we observed strong correlations between fluid intelligence and reasoning for both subtypes (rg_s1_ = −0.23, p_s1_ = 5.87E-06, and rg_s2_ = −0.16, p_s2_ = 0.00020), along with a significant correlation between numeric memory and S1 (rg_s1_ = −0.23, p_s1_ = 0.013) (Supplementary Fig. 36). Moreover, in terms of IDPs from brain and heart, we also observed significant genetic correlations between S1 and RV end-diastolic volume (RVDEV, rg_s1_ = −0.30, p_s1_ = 0.0002), and RV end-systolic volume (RVESV, rg_s1_ = −0.24, p_s1_ = 0.0011) (Supplementary Data. 14). Likewise, associations were noted between S2 and LV myocardial-wall thickness AHA9 (rg_s2_ = 0.20, p_s2_ = 0.0011) and AHA10 (rg_s2_ = 0.24, p_s2_ = 7.33E-05) (Supplementary Data. 14).

Next, we identified the shared causal variant between the two subtypes and outcomes via Bayesian colocalization analyses. Evidence of colocalization was defined as having a posterior probability of the shared causal variant hypothesis (PPH4) > 0.75. Our results revealed that S1 has significant colocalization with bipolar disorder at SNP rs9834970 (Fig. 5c, PPH4 = 0.89) and with chronic kidney disease at SNP rs77924615 (Fig. 5d, PPH4 = 1.0). Moreover, we identified significant colocalization for S2 with asthma at SNP rs7903146 (Fig. 5e, PPH4 = 1.0). These findings underscore both the shared genetic predisposition between the two subtypes and the existence of distinct genetic association contributing to the intricate relationship between subtypes and these outcomes. These findings from genetic associations further indicate the genetic distinctions between the subtypes in relation to different diseases, highlighting the importance of considering subtype-specific genetic profiles in understanding the pathogenesis for these complex conditions.

### Mendelian Randomization for the preclinical-T2DM subtypes with disease outcomes

In light of the robust associations uncovered through phenotypic and genetic analyses, we expanded our inquiry using two-sample MR analyses to explore the underlying causal link between preclinical-T2DM subtypes and disease outcomes. By leveraging instrumental variables (IVs) derived from genome-wide association study (GWAS) summaries of the two subtypes, we identified 2 IVs for S1 and 18 IVs for S2 (Supplementary Tables 8-9). Notably, a lead SNP, rs780094 on chromosome 2 for S2 was excluded due to its strong association with alcohol consumption ^38^, which could potentially be a confounding factor for the causal interpretation between the subtypes and disease outcomes. Following correction for multiple testing using an FDR threshold of p < 0.05, we discerned significant causal relationships for both subtypes with stroke, cardiac arrhythmias, chronic kidney disease, ischemic heart disease, and diabetic retinopathy (Fig. 5f). Notably, several types of MR analysis support the causal association between S2 and these diseases above, while only one MR analysis, IVW, for S1, indicating there still were some differences between the causal relationship identified.

Additionally, we conducted an MR Egger intercept test for S2 to assess the presence of horizontal pleiotropy as applicable, which could potentially bias the causal estimates. The results of the test did not indicate significant horizontal pleiotropy (MR Egger p > 0.05), suggesting that the MR findings for S2 are robust and unbiased (Supplementary Data 15).

### Disease progression prediction using the identified preclinical-T2DM subtypes

Finally, we utilized survival analysis to assess the progression of various diseases in relation to the two preclinical-T2DM subtypes. Survival curves illustrated significant differences in disease progression relative to each subtype (Fig. 6). Subtype 1 exhibited a faster progression and higher risk (higher hazard ratios, HR_S1_ > HR_S2_) for anxiety disorder, bipolar disorder, depression disorder, sleep disorder, obesity, and asthma compared to Subtype 2 (Figs. 6a-f and Supplementary Fig. 48). Conversely, Subtype 2 was associated with more severe progression in Alzheimer’s disease, Parkinson’s disease, hypertension, cardiac arrhythmias, diabetic retinopathy, and a faster progression to T2DM (Figs. 6g-l and Supplementary Fig. 48). These findings align closely with the phenotypic associations previously identified, further revealing the distinct pathophysiological trajectories of each subtype. Of note, the three survival curves of S1, S2, and healthy controls exhibited an initial rapid decline followed by a slowdown towards the end of the follow-up period concerning most comorbidities (refer to Fig. 6 and Supplementary Fig. 48). This observation is primarily attributed to the rapid increase in censoring events, suggesting that the observed effects become less pronounced.

**Fig. 6:**
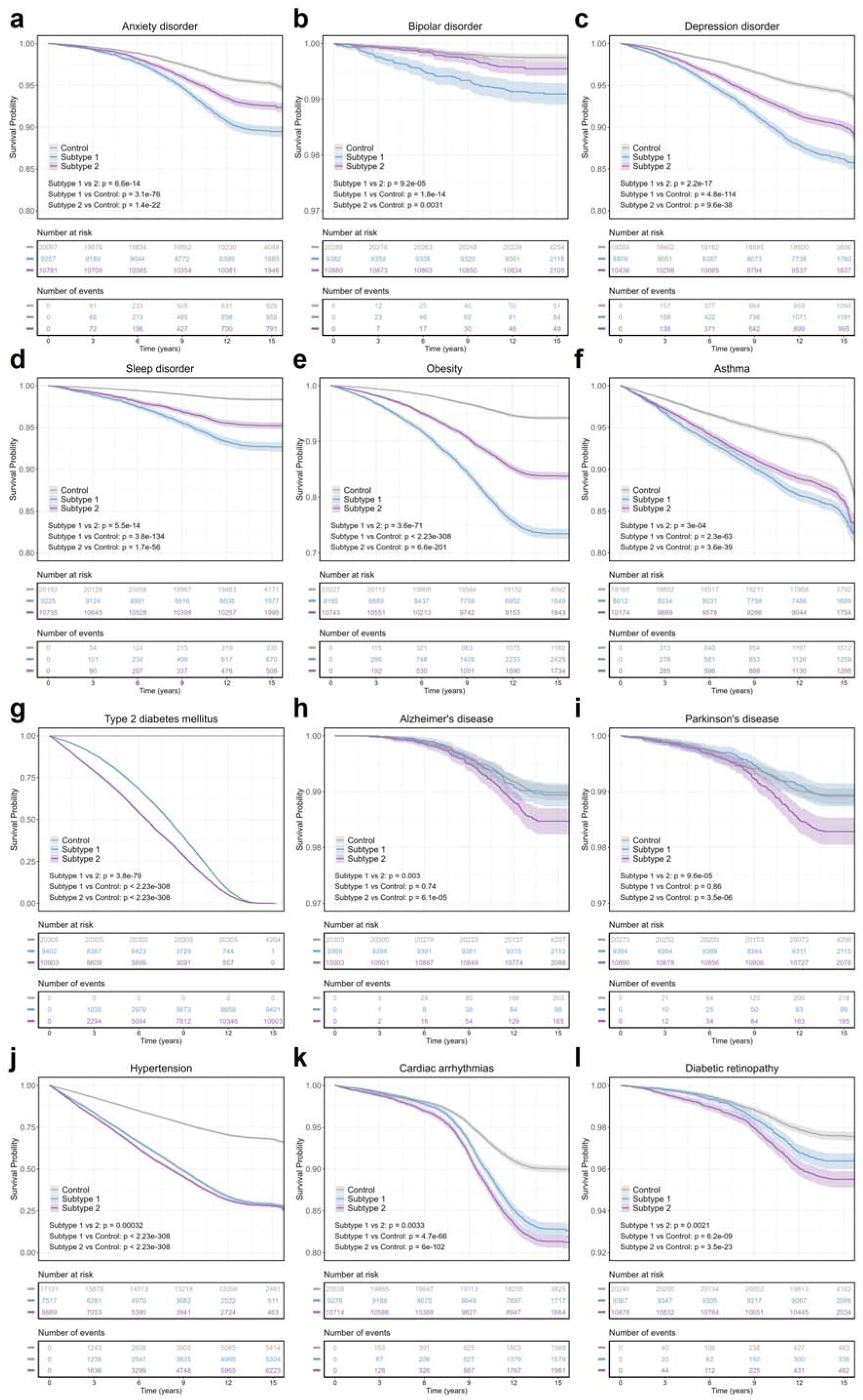
Survival Curves for diseases progression grouping by two preclinical-T2DM subtypes. We illustrated 12 selected diseases that demonstrated significantly distinct progression rates over the years among the three groups, S1, S2, and the control group (log-rank p-value < 0.05). Survival Curves of other diseases were illustrated in Supplementary Figs 37-47. **a-f**, Diseases that showed a faster progression rate in S1 compared to S2 and the control group. **g-l**, Diseases that showed a faster progression rate in S2 compared to S1 and the control group. The two tables below the survival curves present the number of subjects currently at risk (not progressed to the specific disease or censored) and the cumulative number of subjects who had an event (progressed to the specific disease) for each group, corresponding to the years on the x-axis.

Moreover, we evaluated whether the preclinical-T2DM subtypes as biomarkers could enhance the accuracy of disease progression predictions. A baseline model using 18 clinical indicators was enhanced by incorporating information specific to each subtype. The predictive performance was evaluated using concordance index (C-Index). Our results indicated that the inclusion of subtype information could enhance the prediction accuracy for both disease onset and its complications (Supplementary Fig. 49).

## Discussion

In this study, we uncovered the distinct subtypes and stages of preclinical-T2DM by using a cohort of 20,305 UK Biobank participants. Utilizing the machine learning algorithm SuStaIn ^37–39^, we demonstrate that the heterogeneous progression of preclinical-T2DM can be delineated by two distinct trajectories. Both subtypes exhibit different illness durations, biomarker profiles, clinical outcomes and the brain and the heart signatures.

S1, characterized by a higher leptin and lower leptin receptor phenotype, demonstrates elevated levels of BMI, ALT, LDLc, and CRP. S1 has highest circulating leptin and lowest circulating leptin receptor among the healthy control and participants with preclinical-T2DM (Figs. 3g-h). Subjects with leptin resistance have also slightly improved LDL level in the blood (Fig. 2h), which is revealed in S1. It is worth noting that leptin-deficiency is the main cause of massive obesity because of both hyperphagia and decreased energy expenditure ^39–41^. Leptin plays a crucial role in regulating insulin synthesis and secretion from pancreatic β-cells, thereby influencing insulin sensitivity, hepatic glucose production, and glucagon levels ^42,43^. Previous studies have indicated a correlation between leptin resistance and obesity, abnormal cholesterol levels, and heightened risks of psychiatric disorders ^44–46^. Our findings not only confirm these associations but also shed light on the pivotal role of leptin in the initiation and advancement of T2DM. On the contrary, S2 is represented with higher levels of HbA1c and Glucose (Fig. 2d and Table 1). Notably, S2 exhibits a higher proportion of males and a shorter time interval to the onset of T2DM compared to S1 (Table 1). Additionally, the biomarker trajectories of both subtypes differ from each other (Figs. 2e-j, and Supplementary Figs. 5-16). For instance, Glucose and HbA1c levels escalate more rapidly for S2 than for S1 over time (Figs. 2e-f), whereas BMI levels increase more swiftly for S1 compared to S2 during the progression of both subtypes (Fig 2g). These may be caused by a reduced capacity of insulin utilization for participants with S1. However, it is intriguing to note that the progression curve of TyG of S1 initially exceeds that of S2, but the latter rises rapidly and eventually surpasses the former during the progression of preclinical-T2DM (Fig. 2i). This suggests that S1 may not represent a typical insulin-resistant form of T2DM. From a genetic standpoint, S2 manifests more significant genetic associations than S1, with noticeably stronger signals (Fig. 5a). Several identified genetic loci, such as GCKR and IGFI, have been identified as associated with insulin insensitivity ^47,48^. These findings suggest that while S2 is a T2DM genetic-relevant subtype, S1 is more complex and is based on multiple traits that may share common upstream clinical determinants, e.g., leptin resistance. These findings demonstrate the biological plausibility of distinct subtypes of preclinical-T2DM.

We found preclinical-T2DM influences the structures and functions of human organs, particularly brain and heart health. The cumulative damage associated with diabetes can affect multiple organs, including the brain and heart, and we propose that the early susceptibility of neurobiological and cardiovascular structures to metabolic stresses may facilitate the early onset of these damages. Consequently, these structural changes in organs may result in functional impairments. To quantify the associations between preclinical-T2DM and heart and brain health, we analysed brain MRI data from 1,143 individuals and CMR data from 855 individuals in the UK Biobank. After adjusting for various clinical indices, demographics, and imaging confounders, we observed that both subtypes of preclinical-T2DM have long-term effects on many common brain regions and CMR traits, although their impact differs slightly. For example, CMR traits affected by both subtypes, such as increased LV and RV ejection fraction and decreased ascending aorta distensibility (Fig. 4e-f), were associated with a higher risk of cerebrovascular diseases ^49,50^. In addition, a few distinct brain regions and CMR traits were remarkably affected by each subtype. For example, participants with S1 exhibited significant atrophy in the hippocampus and thalamus, while those with S2 showed more pronounced atrophy in the paracentral, left fusiform, and superior frontal regions (Fig. 4a-d). Friedman stated that leptin primarily acts on neurons in the hypothalamus, where it regulates feeding and many other functions, including glucose metabolism ^20^. Previous studies have indicated that “obesity-induced leptin resistance” could be caused by the decreased responsiveness of the hypothalamus to leptin ^51,52^. The reduction of hippocampus, thalamus as well as putamen regions for S1 will predict worse clinical symptoms of mental health, including emotional regulation, stress control, and cognitive performance, which were associated with higher risk of anxiety disorder and depression disorder ^53–56^. For participants with S2, atrophy in paracentral, superior frontal regions were associated with lower limb movement and cognitive impairment in working memory ^57,58^. Notably, the fusiform plays a critical role in cognitive decline associated with language comprehension ^59^, visual processing ^60,61^, and long-term memory ^62^. Atrophy in this region could contribute to Alzheimer’s disease and semantic dementia ^63,64^. Specific associations were also observed between subtypes and CMR traits. For example, the LV circumferential strain traits were decreased in S1. This may suggest impaired myocardial contractility and could be a sign of underlying myocardial damage or early myocardial disease ^65^. A reduction in myocardial wall thickness, as observed in S2 in specific LV segments, are associated with a higher risk of ischemic heart diseases ^66^. Our findings suggest that structural changes in the brain and heart could potentially increase the risk of brain disorders and cardiac diseases. Overall, our study discloses associations between preclinical-T2DM and imaging phenotypes of the brain and heart, indicating potential connections between preclinical-T2DM and cardiovascular and neurological health. While our study specifically focuses on the connections between preclinical-T2DM and the brain/heart, it is also possible to explore the relationships between preclinical-T2DM and other human organs and systems. For example, participants with preclinical-T2DM have a higher risk of chronic kidney disease compared to healthy participants (Fig. 3a). Numerous evidences suggest the interplay among T2DM risk factors, chronic kidney disease, and the cardiovascular system, which has profound impacts on morbidity and mortality. Conducting a multisystem analysis using biobank-scale data may provide insights into interorgan pathophysiological mechanisms and assist in the prevention of T2DM and the early detection of its effects on human organs.

It is widely recognized that individuals exhibit worsening symptoms and face an increased risk of T2DM onset and its complications over the course of the illness. Of note, the observed increases in glucose and HbA1C levels are consistent with an increased risk of stroke, heart failure, ischemic heart disease, chronic kidney disease, etc. These findings align with existing knowledge regarding the crucial role of glucose in cardiovascular and cerebrovascular functions and its causal link with elevated risks of vascular diseases. In addition, both subtypes are correlated with cognitive function decline, a trend mirrored in alterations to brain structure (Figs. 4a-d). Studies have indicated that the probability of T2DM patients experiencing a decline in cognitive abilities is 1.5 to 2.0 times higher than that of non-diabetic individuals ^67–69^. However, despite the occurrence of cognitive function impairment during the preclinical stage of T2DM and the irreversible damage to the brain upon diabetes progression, the adverse effects of preclinical-T2DM on cognitive function are often underestimated, even when clinical symptoms are mild or asymptomatic. Furthermore, it is intriguing to note that both subtypes show differential risks of T2DM and its complications (Fig. 3a and Fig. 6, and Supplementary Fig. 49). For instance, participants with S1 exhibit higher hazard ratios (HRs) than those with S2 for psychiatric disorders, including depression disorder, anxiety disorder, and bipolar disorder, etc. It supports the emerging notion that leptin residence may serve as a potential indicator of neurotransmitter alterations, subsequently impacting the psychiatric status of individuals with preclinical-T2DM S1. Intriguingly, colocalization analysis reveals that genes involved in loci associated with S1 are correlated with bipolar disorder (Fig. 5c). These findings may shed light on why metabolic exposure generally accelerates brain tissue loss in conditions such as bipolar disorder, depression disorder, or other psychiatric disorders, despite being a complex neurobiological process. Conversely, participants with S2 are associated with a higher risk of neurodegenerative disorders, such as Alzheimer’s disease and Parkinson’s disease. Over the past decade, accumulating evidence suggests a positive association between T2DM and dementia ^70–73^. However, our results indicate that this link substantially exists within a subgroup of preclinical-T2DM patients (S2). For individuals with S1, there are no discernible differences between them and healthy controls regarding the risks of Alzheimer’s disease and Parkinson’s disease (Figs. 6h-i). Although further MR analyses suggest no causal associations between Alzheimer’s disease/Parkinson’s disease and preclinical-T2DM (Fig. 5f), our efforts to identify disease subtypes and their associations with brain disorders were conceptual and predominantly relied on accumulating evidence regarding the progression of preclinical-T2DM and its impacts on the brain, which share biological mechanisms with brain disorders. This could potentially aid in the development of cost-effective health promotion strategies tailored to this extensive and vulnerable population.

The identification of distinct trajectories of preclinical-T2DM opens avenues for personalized disease screening and prevention, ultimately leading to improved patient care and outcomes. It is crucial to acknowledge that the subtypes and stages we identified help delineate the heterogeneity of preclinical-T2DM and link them to specific treatments, suggesting that predicting clinical outcomes could benefit from stratification based on the biological subtypes of preclinical-T2DM. To this end, the development of a classifier cluster comprising specific subgroups corresponding to each subtype demonstrates enhanced performance in predicting the onset of T2DM and its complications compared to the model based solely on clinical information. Each subtype exhibits unique clinical characteristics and impacts on human organs, underscoring the importance of tailored approaches in disease management. Previous studies have also suggested the potential benefits of disease risk prediction for certain phenotypes of patients based on specific phenotypic or genetic features ^9,74,75^. While further investigation is warranted on the biological mechanisms of the progression of preclinical-T2DM from the interorgan perspective, factors such as increases in HbA1c and glucose levels, and leptin resistance have consistently shown associations with the disease onset and clinical outcomes. Embracing a perspective of stratified prediction models may unveil the underlying progressive heterogeneity of the disease and facilitate the adoption of more individualized treatment approaches in clinical practice, which holds promise for optimizing patient care, enhancing treatment efficacy, and ultimately mitigating the burden of T2DM and its complications on individuals and healthcare systems alike.

The potential clinical impact of our study is multifaceted. Broadly, it aids in dissecting the heterogeneity of preclinical-T2DM into more defined metabolic subtypes, thereby with implications for downstream tasks. For instance, establishing robust preclinical-T2DM subtypes can enhance the accuracy of individualized disease diagnosis and prognosis. Furthermore, modelling preclinical-T2DM heterogeneity offers novel patient stratification and treatment assessment tools for future clinical trials, which are particularly crucial given the mixed results and clinical limitations of glucose treatments. Recognizing that assessing treatment responses within more homogeneous patient subgroups can significantly enhance the efficacy of clinical trials, our findings suggest that preclinical-T2DM subtyping and staging could improve the ability to identify significant clinical characteristics of T2DM and its complications, which might otherwise be diluted in case-control comparisons due to underlying heterogeneity. Lastly, the identified subtypes, being both phenotypically and genetically relevant to human organs, serve as reliable prognostic biomarkers, thereby facilitating the prediction of disease onset and complication risks.

There are strengths as well as limitations to this study. Firstly, while the SuStaIn algorithm offered estimates of preclinical-T2DM trajectories based on cross-sectional clinical indexes, it is crucial to validate these findings using longitudinal data to authenticate the disease progressions over time. Secondly, we identified two distinct subtypes from the UK Biobank dataset, each exhibiting different clinical phenotypes, neuroanatomical signatures, CMR traits, and clinical outcomes. Validation using independent discovery and replication cohorts would bolster the reliability of these identified subtypes. With the availability of large-scale non-European ancestry populations in studies, there is potential to replicate and validate our findings, particularly in contextualizing the proposed subtypes with brain/heart connectivity, cytoarchitecture, metabolism, neurotransmitter receptors and transporters, gene expression, and cognitive function. Thirdly, while clinical biomarkers and genetics can influence the progression of individuals within preclinical stage of T2DM, the risk factors characterizing the clusters identified in our study are also shaped by behavioural, environmental, and dietary determinants, as well as the use or non-use of medications that lower risk factor levels. Future research integrating these determinants with clinical data is necessary to understand their contributions to the prevalence and trends in preclinical-T2DM subtypes and their impact on disease occurrence.

In summary, we delineate two distinct and stable preclinical-T2DM trajectories originating from cross-sectional clinical data with 18 clinical indexes. These identified subtypes exhibit diverse progressive patterns, clinical symptom profiles, genetic characteristics and long-term outcomes. Our findings highlight the presence of subtypes in preclinical-T2DM and emphasize their importance for personalized treatment and prognostic assessment. This comprehensive understanding of preclinical-T2DM subtypes provides a groundwork for more tailored and effective management strategies, ultimately improving patient outcomes.

## Methods

### Study cohort

In this study, we leveraged the UK Biobank (UKB) dataset, a large biomedical cohort consists of over 500,000 participants as the primary data for our analyses. The use of UKB data has been approved by UKB under application number: 85757. Approval for the UKB study was obtained from the National Research Ethics Committee (REC reference 11/NW/0382), and informed consent was obtained from all participants. The inclusion and exclusion criteria process depicted in Fig. 1a involves the identification of preclinical-T2DM cases and control participants from the UKB dataset. The preclinical-T2DM subjects included in this study were participants with T2DM diagnosis after their initial assessment at the UKB (instance = 0). The UKB dataset comprised 41,783 patients diagnosed with T2DM and 460,628 control participants without T2DM. Diagnoses for T2DM were identified through the International Classification of Diseases, Tenth Revision (ICD-10) codes (E11 for T2DM), as well as self-reported non-cancer T2DM diagnoses. To refine the study population, several exclusion criteria (Fig.1a) were applied to excluded individuals who: (1) diagnosed with other forms of diabetes; (2) were on antidiabetic medications or had history of antidiabetic medication usage; (3) lacked crucial genomic data for subsequent genetic association studies; (4) with outlier biomarker values that deviating more than five standard deviations from the mean; (5) were diagnosed with T2DM prior to their initial assessment at the UKB to focus on preclinical cases; (6) were potential undiagnosed diabetes in the control group with HbA1c levels equal to or greater than 6.5% (47 mmol/mol), or glucose levels equal to or greater than 7 mmol/L. This exclusion process yields a total of 20,305 individuals (aged 40-70 years, mean age 59.60 years, 42.28% female) with preclinical-T2DM.

Lastly, we employed propensity score matching (PSM) for the construction of a matched control group to ensure the computability of the SuStaIn model. PSM is a statistical technique used to reduce bias by balancing the distribution of confounding variables between two groups. Specifically, we applied PSM to harmonize covariates between individuals exhibiting preclinical-T2DM and those in the control group. Covariates such as age, sex, smoking status, alcohol drinking status, income level, and education attainment were incorporated to estimate the probability of each individual being assigned to the preclinical-T2DM group. Then, we matched individuals with similar probabilities from the control group to ensure that the distribution of these covariates was similar between the two groups. Thereafter, we established a balanced control group of 20,305 individuals without any diabetes (aged 40-73 years, mean age 59.96 years, 41.73% female) with the least standard mean difference (SMD) compared with the preclinical-T2DM group for subsequent analyses (Supplementary Table 1).

### Feature selection process

Initially, we collected 62 biomarkers, including blood cell counts, biochemical markers, lipid profiles, blood pressure, and BMI (Supplementary Table 2) from the UKB dataset for SuStaIn modelling. These biomarkers were accessed for each individual at their initial assessment at the UKB. Missing data (all biomarkers with a missing rate < 30%) were imputed using the multivariate imputation by chained equation (MICE) from R package “MICE” (version 3.16). Random forest was implemented as the main method for imputation. The imputation was performed 5 times, with 5 iterations for each imputation, then the average of the 5 times of imputation was taken as the final imputation result. Given the computational demands of the SuStaIn model and insight from prior research suggesting an optimal number of features of approximately 20 for computational feasibility ^8,10^, we implemented a cutoff criterion to select the most important biomarkers for SuStaIn modelling. Specifically, we quantified the influence of each biomarker on preclinical-T2DM through univariate logistic regression analyses to determine their effect size relative to preclinical-T2DM. Biomarkers with an absolute effect size exceeding 0.3 were retained for SuStaIn modelling. This process identified 18 pivotal biomarkers, consisting of hemoglobin A1c (HbA1c), body mass index (BMI), High-density lipoprotein cholesterol (HDLc), triglycerides-glucose index (TyG), high light scatter reticulocyte count (HLS Retic), glucose, reticulocyte count (RET), immature reticulocyte fraction (IRF), apolipoprotein A (ApoA), alanine aminotransferase (ALT), triglycerides (TG), urate, white blood cell count (WBC), cholesterol (CHOL), c-reactive protein (CRP), lymphocyte count (LYM), Low-density lipoprotein cholesterol (LDLc) and Vitamin D. For detailed information on the selected biomarkers, please refer to Supplementary Table 2.

### Identification of subtypes and stages for preclinical-T2DM

To unveil the diverse manifestations of preclinical-T2DM and explore its interplay with the health of the brain, the heart, and other organs related outcomes, we utilized the SuStaIn model to categorize participants with preclinical-T2DM into distinct subtypes and stages of disease progression. SuStaIn, an unsupervised machine-learning technique, identifies clusters that share similar patterns of disease evolution using cross-sectional data using an event-based model ^8,76^. Initially crafted for neurodegenerative conditions like Alzheimer’s disease ^70^, SuStaIn has proven its effectiveness in delineating unique subtypes and their progression in various disorders, including major depressive disorder ^11^ schizophrenia ^12^ and epilepsy ^10,13^. These insights highlight the adaptability of SuStaIn in unveiling disease subtypes and progression stages, which has significant implications for the care and management over the course of T2DM.

We applied the linear z-scored SuStaIn model ^11,77^, integrating the 18 selected clinical biomarkers for subtype and stage identification. Prior to analysis, potential confounding variables, including age, sex, smoking status, alcohol drinking status, income level, and educational attainment, were adjusted for each biomarker. Subsequently, the adjusted biomarkers were z-scored relative to the control group. Model initialization was achieved through an expectation-maximization algorithm, executed across 25 distinct random starting points to determine the maximum likelihood solution. We conducted 100,000 iterations of the Markov Chain Monte Carlo (MCMC) method to estimate uncertainty surrounding the progression patterns of identified subtypes.

Furthermore, to ascertain the optimal number of preclinical-T2DM subtypes, we employed a ten-fold cross-validation approach, ranging from 1 to 5 subtypes. Performance evaluation was conducted using the Cross-Validation Information Criterion (CVIC) ^11,77^. The CVIC is a metric used to evaluate the fit of a model while penalizing the model’s complexity, ensuring that the model with the optimal balance of simplicity and accuracy is selected. We observed that compared to the 2-subtype model, the models with 3-5 subtypes show less than a 2% improvement in CVIC (Supplementary Fig. 1). Considering the 2-subtype model for its optimal balance of simplicity and explanatory power, we chose to interpret the findings based on the 2-subtype model.

### Phenotypic associations between preclinical-T2DM subtypes and outcomes

We examined the phenotypic associations between preclinical-T2DM subtypes and a various range of health outcomes. Our analysis included a broad spectrum of phenotypic measures, consisting of organ structure and function of brain and heart, diseases including brain, cardiac, and renal disorders, as well as other related disorders, cognitive functions, metabolic and proteomic profiles.

For brain structure and function, we analysed 587 image-derived phenotypes (IDPs) extracted from T1-weighted magnetic resonance imaging (MRI) and diffusion tensor imaging (DTI), as outlined by Guo et al ^78^. Image protocols and pre-processing pipelines of the brain MRIs were sourced from the UKB in an online document (https://biobank.ctsu.ox.ac.uk/crystal/crystal/docs/brain_mri.pdf). Data of Brain IDPs were available for 1134 preclinical-T2DM subjects, with 520 for S1, and 614 for S2, respectively. Specifically, the 587 IDPs consist of 203 cortical measurements from the FreeSurfer tool ^79^ based on the Desikan-Killiany atlas ^80^, 24 subcortical measurements extracted by the FIRST tool ^81^, and the FreeSurfer tool ^81^ based on automatic subcortical segmentation ^82^, along with 360 IDPs of white matter connections derived from two complementary analyses: tract-based spatial statistics ^83^ based on the International Consortium of Brain Mapping (ICBM) DTI-81 atlas ^84^, and probabilistic tractography ^85^.

Cardiac and aortic functions were evaluated using 82 imaging traits for left ventricle (LV), left atrium (LA), right ventricle (RV), right atrium (RA), aorta (Ao), ascending aorta, and descending aorta, obtained from recently developed pipelines for cardiac and aortic MRI ^74,86,87^. The CMR traits were available for 855 preclinical-T2DM subjects with 380 subjects for S1, and 475 subjects for S2, respectively.

The prevalence of various disorders was identified using ICD-10 codes and self-reported non-cancer diagnoses from the UKB, as detailed in Supplementary Table 3. Additionally, cognitive function was assessed across several aspects provided by UKB, including numeric memory, symbol digit substitution, reaction time, fluid intelligence and reasoning, prospective memory, and trail making (Supplementary Table 4). Moreover, the correspondent proteomic and metabolic data at baseline for the studied cohort were obtained from UKB, including 1463 proteomic biomarkers, and 251 metabolic indictors (Supplementary Data 7-8).

To assess the phenotypic associations, we employed univariate regression analyses to determine the impacts of different preclinical-T2DM subtypes compared to the control group on organ structural and functional variables as well as cognitive functions. Covariates including sex, age, smoking status, alcohol drinking status, income level, and education attainment were adjusted to control for potential confounding effects. Furthermore, the odds ratios (ORs) for the incidence of various diseases across the two preclinical-T2DM subtypes were calculated to measure the relative risk conferred by each subtype. The statistical significance of differences observed between the subtypes and control groups regarding disease occurrence was evaluated using the *t*-test. Moreover, we explored the difference in the proteomic and metabolomic profiles between the two preclinical-T2DM subtypes and healthy controls. For the proteomic and metabolic data, Student’s *t*-test with equal variance was applied in the differential analysis between the two groups. Additionally, one-way ANOVA with equal variance was utilized for the differential analysis encompassing all three groups. Significant associations were identified based on a false discovery rate (FDR) threshold of 5%, which was adjusted for multiple testing using the Benjamini-Hochberg procedure.

### GWAS on preclinical-T2DM subtypes

We conducted genome-wide association studies (GWAS) using genotyped and imputed data from the UKB on both subtypes of preclinical-T2DM. Genome-wide genotyping data was performed on all UKB participants using the UK Biobank Axiom Array, followed by imputation using the Haplotype Reference Consortium and UK10K as reference panels (GRCh37 assembly) ^88^. The analyses were stratified into two comparative sets: S1 (n = 9,402) vs controls (n = 20,305) and S2 (n = 10,903) vs the same control group (n = 20,305). Before conducting GWAS analyses, stringent quality control measures were implemented. Specifically, samples exhibiting a missingness rate greater than 0.05, minor allele frequency (MAF) less than 0.01, or deviation from Hardy-Weinberg Equilibrium (HWE) with a p-value less than 1.0E-06 were excluded from the analyses. The GWAS analyses were performed using the PLINK software (version 1.90 beta, https://www.cog-genomics.org/plink/, with adjustments for covariates including sex, age, smoking status, alcohol drinking status, income level, education attainment, and the first ten principal components to address potential population stratification effects. The genome-wide significance threshold was set at 5.0E-08. We employed the Functional Mapping and Annotation (FUMA) ^89^ platform (https://fuma.ctglab.nl/) with default settings to identify and annotate lead SNPs and corresponding genes, within a slide window size of 500 kilobases (kb) of the significant loci. The 1000 Genomes Project Phase 3 European (1KGp3 EUR) panel were employed as the reference dataset. SNPs with the smallest association p-values were selected as the lead SNPs, and genes residing within the boundaries of each region were allocated for each of the identified risk loci.

### Genetic correlation and colocalization between preclinical-T2DM subtypes and outcomes

To evaluate the genetic associations between preclinical-T2DM subtypes and outcomes, we employed two genetic analysis techniques: genetic correlation and colocalization analysis, utilizing publicly available GWAS results primarily obtained from FinnGen database (https://www.finngen.fi/), Psychiatric Genomics Consortium (PGC, https://med.unc.edu/pgc/download-results) and GWAS Catalog (https://www.ebi.ac.uk/gwas/). Detailed information on the GWAS studies and results for the outcomes utilized for this analysis can be found in Supplementary Table 7. Firstly, we utilized linkage disequilibrium score regression (LDSC, https://github.com/bulik/ldsc) ^90^ to measure the genetic correlation between the T2DM subtypes and outcomes. Only high-quality SNPs documented in the HapMap3 dataset were utilized for estimation, with the LD score derived from the 1KGp3 EUR panel employed for LDSC analysis.

Furthermore, to determine whether the preclinical-T2DM subtypes share a common causal variant with the outcomes, we conducted colocalization analysis using the R package “coloc” (https://chr1swallace.github.io/coloc/index.html) ^91–93^. This method employs Bayesian statistics to estimate the posterior probabilities of five distinct hypotheses concerning the relationship between the association signals at a shared locus: PPH0 (No association with either trait), H1 (Association with the first trait only), PPH2 (Association with the second trait only), PPH3 (Associations with both traits, but with different causal variants), and PPH4 (Association with both traits due to the same causal variant). Colocalization was performed using with default priors (prior probability of initial trait association is 1.0E-04, prior probability of shared causal variant across two traits is 1.0E-05). In accordance with established conventions ^94^, variants with a posterior probability of PPH4 > 0.75 were considered as colocalized variants (indicating shared causal variants) for preclinical-T2DM subtypes and outcomes.

### Mendelian Randomization analyses

To further explore the causal relationship between preclinical-T2DM subtypes and outcomes, we conducted two-sample Mendelian Randomization (MR) analyses using the R package “TwoSampleMR” (https://mrcieu.github.io/TwoSampleMR/). SNPs that met the significance threshold (p < 5.0 × 10^-8^) were selected as candidate instrumental variables (IVs). Next, independent SNPs were chosen from GWAS summary data of the preclinical-T2DM subtypes utilizing the clump function in Plink. These selected SNPs were clumped at an r^2^ = 0.1 within a 1000 kb window size, employing the LD score panel of 1KGp3 EUR to adjust for linkage disequilibrium and reduce interference. Furthermore, SNPs strongly associated with confounders (p < 5.0E-08) that might affect the pathway between subtype and outcome causation were excluded. We considered five potential confounders: smoking status, alcohol drinking status, income level, educational attainment, and BMI. Additionally, to enhance the robustness of the genetic instruments, we assessed the *F*-statistic to measure the statistical power of the IVs and removed weak instruments with *F*-statistic < 10. Consequently, this IV selection process resulted in 2 IVs for S1 and 18 IVs for S2.

For S1, we performed MR analyses using the inverse variance weighted (IVW) method due to the limited availability of only 2 IVs. For S2, we employed four additional MR methods: MR Egger, MR-RAPS, weighted median, and weighted mode. This aimed to enhance the reliability of the results, as IVW results might be biased in the presence of horizontal pleiotropy in any of the SNPs. We regarded significant findings as those surpassing the threshold adjusted using Benjamini-Hochberg procedure to control the FDR at the 5% level (FDR-corrected p-value < 0.05). Furthermore, we assessed the potential presence of horizontal pleiotropy, which could compromise the validity of the MR findings. We conducted MR-Egger intercept test to detect the presence of horizontal pleiotropy for MR findings for S2 with three or more IVs available. A p-value greater than 0.05 indicates nonsignificant presence of horizontal pleiotropy.

### Prediction analyses

To evaluate the predictive power of these subtypes for disease progression, we examined the efficacy and applicability of preclinical-T2DM subtypes as predictive clinical indicators for various diseases as outcomes. Firstly, we analysed survival curves for different diseases across the subtypes to assess the rate of disease progression associated with each subtype. Individuals not diagnosed with the disease by the end of the observation period were considered as censoring. The censoring date was set at the latest access date (May 31, 2023) for the UK Biobank data. We utilized the log-rank test to assess differences in survival distributions between each subtype and the control group as the reference. Secondly, we employed Cox regression models to estimate hazard ratios (HRs) for the onset of each of diseases, comparing the two subtypes.

We further investigated whether incorporating subtype information could enhance disease prediction. Cox regression models were utilized, integrating different subtype information as predictors. A baseline model was first established using 18 clinical biomarkers, with subsequent integration of different subtypes to assess their impact on predictive accuracy. To validate the robustness of our findings, we employed a ten-fold cross-validation method. The dataset was equally divided into ten parts, with nine parts used as the training set and the remaining part used as the test set in each validation cycle. The concordance index (C-index) was utilized as the primary metric to evaluate the predictive performance of the models. The C-index measures the predictive accuracy of the model in terms of its ability to correctly rank the survival times of subjects, with higher values indicating better predictive accuracy.

## Data availability

The data used and generated in this study are provided in the Supplementary Data. The phenotypic, genotypic, proteomic, metabolic data used in the study that supports subtype and stage modelling and association analyses were obtained from the UK Biobank under application number 89757. Access to the UK Biobank data is available to all researchers with approval (https://www.ukbiobank.ac.uk/enable-your-research/register). All GWAS data for the outcomes used in this study are publicly available from FinnGen database (https://www.finngen.fi/), Psychiatric Genomics Consortium (PGC, https://med.unc.edu/pgc/download-results) and GWAS Catalog (https://www.ebi.ac.uk/gwas/). Statistic details of the GWAS datasets and download links for all the datasets are available in Supplementary Table 7.

## Code availability

The original source code for the implementation of SuStaIn algorithm is available on the GitHub repository at https://github.com/ucl-pond/pySuStaIn. The source codes pertaining to this study and data analysis in this manuscript are provided at https://github.com/ZJU-BMI/disease-progression-preclinical-T2DM.

## Author contributions

ZH and FW jointly supervised research. ZH designed this study. FY developed a deep learning model and performed the model interpretation. FY, ZJ and ZH performed data analysis. FY, JY, FH, MP, JS, ZJ, FW, YZ and ZH interpreted the results. FY, ZJ, MP, JS, YZ and ZH prepared the first draft of the manuscript. All authors contributed and approved the final draft.

## Acknowledgements

The UK Biobank resource was used under application number 89757.

## References

1. Sun, H. et al. IDF Diabetes Atlas: Global, regional and country-level diabetes prevalence estimates for 2021 and projections for 2045. Diabetes Res. Clin. Pract. 183, 109119 (2022).

2. Kumar, A., Gangwar, R., Ahmad Zargar, A., Kumar, R. & Sharma, A. Prevalence of diabetes in India: A review of IDF Diabetes Atlas 10th edition. Curr. Diabetes Rev. (2023) doi:10.2174/1573399819666230413094200.

3. Galicia-Garcia, U. et al. Pathophysiology of type 2 Diabetes Mellitus. Int. J. Mol. Sci. 21, 6275 27 (2020).

4. Garcia-Serrano, A. M. & Duarte, J. M. N. Brain metabolism alterations in type 2 diabetes: What did we learn from diet-induced diabetes models? Front. Neurosci. 14, 229 (2020).

5. Xourafa, G., Korbmacher, M. & Roden, M. Inter-organ crosstalk during development and progression of type 2 diabetes mellitus. Nat. Rev. Endocrinol. (2023) doi:10.1038/s41574-023-00898-1.

6. Zheng, R. et al. Data-driven subgroups of prediabetes and the associations with outcomes in Chinese adults. Cell Rep. Med. 4, 100958 (2023).

7. Young, A. L. et al. Data-driven modelling of neurodegenerative disease progression: thinking outside the black box. Nat. Rev. Neurosci. (2024) doi:10.1038/s41583-023-00779-6.

8. Young, A. L. et al. Uncovering the heterogeneity and temporal complexity of neurodegenerative diseases with Subtype and Stage Inference. Nat. Commun. 9, 4273 (2018).

9. Tao, P. et al. Metabolomics and Lipidomics Analyses Aid Model Classification of Type 2 Diabetes in Non-Human Primates. Metabolites 14, (2024).

10. Jiang, Y. et al. Identification of four biotypes in temporal lobe epilepsy via machine learning on brain images. Nat. Commun. 15, 2221 (2024).

11. Chen, D. et al. Neurophysiological stratification of major depressive disorder by distinct trajectories. Nature Mental Health 1, 863–875 (2023).

12. Jiang, Y. et al. Neuroimaging biomarkers define neurophysiological subtypes with distinct trajectories in schizophrenia. Nature Mental Health 1, 186–199 (2023).

13. Xiao, F. et al. Identification of different MRI atrophy progression trajectories in epilepsy by subtype and stage inference. Brain 146, 4702–4716 (2023).

14. Cholerton, B., Baker, L. D., Montine, T. J. & Craft, S. Type 2 diabetes, cognition, and dementia in older adults: Toward a precision health approach. Diabetes Spectr. 29, 210–219 (2016).

15. Zhang, Y. et al. Effects of vitamin D supplementation on prevention of type 2 diabetes in patients with prediabetes: A systematic review and meta-analysis. Diabetes Care 43, 1650–1658 (2020).

16. Geng, L., Lam, K. S. L. & Xu, A. The therapeutic potential of FGF21 in metabolic diseases: from bench to clinic. Nat. Rev. Endocrinol. 16, 654–667 (2020).

17. Suriano, F. et al. Novel insights into the genetically obese (ob/ob) and diabetic (db/db) mice: two sides of the same coin. Microbiome 9, 147 (2021).

18. Bungau, S. et al. Interactions between leptin and insulin resistance in patients with prediabetes, with and without NAFLD. Exp. Ther. Med. 20, 197 (2020).

19. Mazor, R. et al. Cleavage of the leptin receptor by matrix metalloproteinase-2 promotes leptin resistance and obesity in mice. Sci. Transl. Med. 10, eaah6324 (2018).

20. Friedman, J. M. Leptin and the endocrine control of energy balance. Nat. Metab. 1, 754–764 (2019).

21. Rajwani, A. et al. Increasing circulating IGFBP1 levels improves insulin sensitivity, promotes nitric oxide production, lowers blood pressure, and protects against atherosclerosis. Diabetes 61, 915–924 (2012).

22. Herder, C. et al. Genetic determinants of circulating interleukin-1 receptor antagonist levels and their association with glycemic traits. Diabetes 63, 4343–4359 (2014).

23. Tommy H T Wong, Jacky M Y Mo, Mingqi Zhou, Jie V Zhao, C Mary Schooling, Baoting He, Shan Luo & Shiu Lun. A two-sample Mendelian randomization study explores metabolic profiling of different glycemic traits. Commun Biol. 7, 293 (2024).

24. Wittenbecher, C. et al. Insulin-like growth factor binding protein 2 (IGFBP-2) and the risk of developing type 2 diabetes. Diabetes 68, 188–197 (2019).

25. Del Bosque-Plata, L., Martínez-Martínez, E., Espinoza-Camacho, M. Á. & Gragnoli, C. The role of TCF7L2 in type 2 diabetes. Diabetes 70, 1220–1228 (2021).

26. Fernandes Silva, L., Vangipurapu, J., Kuulasmaa, T. & Laakso, M. An intronic variant in the GCKR gene is associated with multiple lipids. Sci. Rep. 9, 10240 (2019).

27. Kovacs, P. et al. The role of insulin receptor substrate-1 gene (IRS1) in type 2 diabetes in Pima Indians. Diabetes 52, 3005–3009 (2003).

28. Mastracci, T. L., Colvin, S. C., Padgett, L. R. & Mirmira, R. G. Hypusinated eIF5A is expressed in the pancreas and spleen of individuals with type 1 and type 2 diabetes. PLoS One 15, e0230627 (2020).

29. Ghosh, C. et al. Involvement of Cdkal1 in the etiology of type 2 diabetes mellitus and microvascular diabetic complications: a review. J. Diabetes Metab. Disord. 21, 991–1001 (2022).

30. Viñuela, A. et al. Genetic variant effects on gene expression in human pancreatic islets and their implications for T2D. Nat. Commun. 11, 4912 (2020).

31. Kobiita, A. et al. The diabetes gene JAZF1 is essential for the homeostatic control of ribosome biogenesis and function in metabolic stress. Cell Rep. 32, 107846 (2020).

32. Flannick, J. et al. Loss-of-function mutations in SLC30A8 protect against type 2 diabetes. Nat. Genet. 46, 357–363 (2014).

33. Zhang, J., McKenna, L. B., Bogue, C. W. & Kaestner, K. H. The diabetes gene Hhex maintains δ-cell differentiation and islet function. Genes Dev. 28, 829–834 (2014).

34. Erfani, T. et al. KCNQ1 common genetic variant and type 2 diabetes mellitus risk. J. Diabetes Metab. Disord. 19, 47–51 (2020).

35. Li, L. et al. The effect of lncRNA-ARAP1-AS2/ARAP1 on high glucose-induced cytoskeleton rearrangement and epithelial-mesenchymal transition in human renal tubular epithelial cells. J. Cell. Physiol. 235, 5787–5795 (2020).

36. Steinthorsdottir, V. et al. Identification of low-frequency and rare sequence variants associated with elevated or reduced risk of type 2 diabetes. Nat. Genet. 46, 294–298 (2014).

37. Mellado-Gil, J. M. et al. The type 2 diabetes-associated HMG20A gene is mandatory for islet beta cell functional maturity. Cell Death Dis. 9, 279 (2018).

38. Jorgenson, E. et al. Genetic contributors to variation in alcohol consumption vary by race/ethnicity in a large multi-ethnic genome-wide association study. Mol. Psychiatry 22, 1359–1367 (2017).

39. Xu, J. et al. Genetic identification of leptin neural circuits in energy and glucose homeostases. Nature 556, 505–509 (2018).

40. Clément, K. et al. Efficacy and safety of setmelanotide, an MC4R agonist, in individuals with severe obesity due to LEPR or POMC deficiency: single-arm, open-label, multicentre, phase 3 trials. Lancet Diabetes Endocrinol. 8, 960–970 (2020).

41. Takahashi, K. et al. Inter-organ insulin-leptin signal crosstalk from the liver enhances survival during food shortages. Cell Rep. 42, 112415 (2023).

42. Pereira, S., Cline, D. L., Glavas, M. M., Covey, S. D. & Kieffer, T. J. Tissue-specific effects of Leptin on glucose and lipid metabolism. Endocr. Rev. 42, 1–28 (2021).

43. Duquenne, M. et al. Leptin brain entry via a tanycytic LepR-EGFR shuttle controls lipid metabolism and pancreas function. Nat. Metab. 3, 1071–1090 (2021).

44. Changchien, T.-C., Tai, C.-M., Huang, C.-K., Chien, C.-C. & Yen, Y.-C. Psychiatric symptoms and leptin in obese patients who were bariatric surgery candidates. Neuropsychiatr. Dis. Treat. 11, 2153–2158 (2015).

45. Dallner, O. S. et al. Dysregulation of a long noncoding RNA reduces leptin leading to a leptin-responsive form of obesity. Nat. Med. 25, 507–516 (2019).

46. Qin Zeng, Jianfeng Song, Xiaoxiao Sun, Dandan Wang, Xiyan Liao, Yujin Ding, Wanyu Hu, Yayi Jiao, Wuqian Mai, Wufuer Aini, Fanqi Wang, Hui Zhou, Limin Xie, Ying Mei, Yuan Tang, Zhiguo Xie, Haijing Wu, Wei Liu & Tuo Deng. A negative feedback loop between TET2 and leptin in adipocyte regulates body weight. Nat Commun. 15, 2825 (2024).

47. Kimura, M. et al. En masse organoid phenotyping informs metabolic-associated genetic susceptibility to NASH. Cell 185, 4216–4232.e16 (2022).

48. Barbieri, M., Bonafè, M., Franceschi, C. & Paolisso, G. Insulin/IGF-I-signaling pathway: an evolutionarily conserved mechanism of longevity from yeast to humans. Am. J. Physiol. Endocrinol. Metab. 285, E1064–71 (2003).

49. Ahmadi, N. et al. Impaired aortic distensibility measured by computed tomography is associated with the severity of coronary artery disease. Int. J. Cardiovasc. Imaging 27, 459–469 (2011).

50. Chue, C. D., Edwards, N. C., Ferro, C. J., Townend, J. N. & Steeds, R. P. Effects of age and chronic kidney disease on regional aortic distensibility: a cardiovascular magnetic resonance study. Int. J. Cardiol. 168, 4249–4254 (2013).

51. Ahima, R. S. & Flier J. S. Leptin. Annu. Rev. Physiol. 62, 413–437 (2000).

52. Friedman, J. The long road to leptin. J. Clin. Invest. 126, 4727–4734 (2016).

53. Talati, A. et al. Putamen structure and function in familial risk for depression: A multimodal imaging study. Biol. Psychiatry 92, 932–941 (2022).

54. Lin, H., Bruchmann, M. & Straube, T. Altered putamen activation for social comparison-related feedback in social anxiety disorder: A pilot study. Neuropsychobiology 82, 359–372 (2023).

55. Anacker, C. et al. Hippocampal neurogenesis confers stress resilience by inhibiting the ventral dentate gyrus. Nature 559, 98–102 (2018).

56. Morelli, M. E. et al. Early putamen hypertrophy and ongoing hippocampus atrophy predict cognitive performance in the first ten years of relapsing-remitting multiple sclerosis. Neurol. Sci. 41, 2893–2904 (2020).

57. Murayama, T., Takahama, K., Jinbo, K. & Kobari, T. Anatomical increased/decreased changes in the brain area following individuals with chronic traumatic complete thoracic spinal cord injury. Phys. Ther. Res. 24, 163–169 (2021).

58. Klingberg, T. Development of a superior frontal-intraparietal network for visuo-spatial working memory. Neuropsychologia 44, 2171–2177 (2006).

59. Qu, J. et al. Cross-language pattern similarity in the bilateral fusiform cortex is associated with reading proficiency in second language. Neuroscience 410, 254–263 (2019).

60. Davies-Thompson, J., Johnston, S., Tashakkor, Y., Pancaroglu, R. & Barton, J. J. S. The relationship between visual word and face processing lateralization in the fusiform gyri: A cross-sectional study. Brain Res. 1644, 88–97 (2016).

61. Schindler, S., Wegrzyn, M., Steppacher, I. & Kissler, J. Perceived communicative context and emotional content amplify visual word processing in the fusiform gyrus. J. Neurosci. 35, 6010–6019 (2015).

62. Zhu, B. et al. Hippocampal size is related to short-term true and false memory, and right fusiform size is related to long-term true and false memory. Brain Struct. Funct. 221, 4045–4057 (2016).

63. Ma, D. et al. The fusiform gyrus exhibits an epigenetic signature for Alzheimer’s disease. Clin. Epigenetics 12, 129 (2020).

64. Landin-Romero, R., Tan, R., Hodges, J. R. & Kumfor, F. An update on semantic dementia: genetics, imaging, and pathology. Alzheimers. Res. Ther. 8, 52 (2016).

65. Marek, J. et al. Three-dimensional echocardiographic left ventricular strain analysis in Fabry disease: correlation with heart failure severity, myocardial scar, and impact on long-term prognosis. Eur. Heart J. Cardiovasc. Imaging 24, 1629–1637 (2023).

66. Liga, R., Colli, A., Taggart, D. P., Boden, W. E. & De Caterina, R. Myocardial revascularization in patients with ischemic cardiomyopathy: For whom and how. J. Am. Heart Assoc. 12, e026943 (2023).

67. Ennis, G. E., Saelzler, U., Umpierrez, G. E. & Moffat, S. D. Prediabetes and working memory in older adults. Brain Neurosci. Adv. 4, 2398212820961725 (2020).

68. Parashar, A., Mehta, V. & Malairaman, U. Type 2 diabetes mellitus is associated with social recognition memory deficit and altered dopaminergic neurotransmission in the amygdala. Ann. Neurosci. 24, 212–220 (2018).

69. Koekkoek, P. S., Kappelle, L. J., van den Berg, E., Rutten, G. E. H. M. & Biessels, G. J. Cognitive function in patients with diabetes mellitus: guidance for daily care. Lancet Neurol. 14, 329–340 (2015).

70. Biessels, G. J. & Despa, F. Cognitive decline and dementia in diabetes mellitus: mechanisms and clinical implications. Nat. Rev. Endocrinol. 14, 591–604 (2018).

71. Ehtewish, H., Arredouani, A. & El-Agnaf, O. Diagnostic, prognostic, and mechanistic biomarkers of diabetes mellitus-associated cognitive decline. Int. J. Mol. Sci. 23, 6144 (2022).

72. Du, H., Meng, X., Yao, Y. & Xu, J. The mechanism and efficacy of GLP-1 receptor agonists in the treatment of Alzheimer’s disease. Front. Endocrinol. (Lausanne) 13, 1033479 (2022).

73. Zhou, Y., Dong, J., Song, J., Lvy, C. & Zhang, Y. Efficacy of glucose metabolism-related indexes on the risk and severity of Alzheimer’s disease: A meta-analysis. J. Alzheimers. Dis. 93, 1291–1306 (2023).

74. Zhao, B. et al. Heart-brain connections: Phenotypic and genetic insights from magnetic resonance images. Science 380, abn6598 (2023).

75. Jingning Zhang, Jianan Zhan, Jin Jin, Cheng Ma, Ruzhang Zhao, Jared O’Connell, Yunxuan Jiang, 23andMe Research Team, Bertram L. Koelsch, Haoyu Zhang & Nilanjan Chatterjee. An ensemble penalized regression method for multi-ancestry polygenic risk prediction. Nat Commun . 15, 3238 (2024).

76. Fonteijn, H. M. et al. An event-based model for disease progression and its application in familial Alzheimer’s disease and Huntington’s disease. Neuroimage 60, 1880–1889 (2012).

77. Aksman, L. M. et al. pySuStaIn: a Python implementation of the Subtype and Stage Inference algorithm. SoftwareX 16, (2021).

78. Guo, J. et al. Mendelian randomization analyses support causal relationships between brain imaging-derived phenotypes and risk of psychiatric disorders. Nat. Neurosci. 25, 1519–1527 (2022).

79. Fischl B. FreeSurfer. Neuroimage 62, 774–781 (2012).

80. Iscan, Z. et al. Test-retest reliability of freesurfer measurements within and between sites: Effects of visual approval process. Hum. Brain Mapp. 36, 3472–3485 (2015).

81. Nugent, A. C. et al. Automated subcortical segmentation using FIRST: test-retest reliability, interscanner reliability, and comparison to manual segmentation. Hum. Brain Mapp. 34, 2313–2329 (2013).

82. Fischl, B. et al. Whole brain segmentation: automated labeling of neuroanatomical structures in the human brain. Neuron 33, 341–355 (2002).

83. Smith, S. M. et al. Tract-based spatial statistics: voxelwise analysis of multi-subject diffusion data. Neuroimage 31, 1487–1505 (2006).

84. Mori, S. et al. Stereotaxic white matter atlas based on diffusion tensor imaging in an ICBM template. Neuroimage 40, 570–582 (2008).

85. Behrens, T. E. J., Berg, H. J., Jbabdi, S., Rushworth, M. F. S. & Woolrich, M. W. Probabilistic diffusion tractography with multiple fibre orientations: What can we gain? Neuroimage 34, 144–155 (2007).

86. Bai, W. et al. A population-based phenome-wide association study of cardiac and aortic structure and function. Nat. Med. 26, 1654–1662 (2020).

87. Bai, W. et al. Recurrent neural networks for aortic image sequence segmentation with sparse annotations. in Medical Image Computing and Computer Assisted Intervention – MICCAI 2018 586–594 (Springer International Publishing, Cham, 2018).

88. Bycroft, C. et al. The UK Biobank resource with deep phenotyping and genomic data. Nature 562, 203–209 (2018).

89. Watanabe, K., Taskesen, E., van Bochoven, A. & Posthuma, D. Functional mapping and annotation of genetic associations with FUMA. Nat. Commun. 8, 1826 (2017).

90. Bulik-Sullivan, B. K. et al. LD Score regression distinguishes confounding from polygenicity in genome-wide association studies. Nat. Genet. 47, 291–295 (2015).

91. Giambartolomei, C. et al. Bayesian test for colocalisation between pairs of genetic association studies using summary statistics. PLoS Genet. 10, e1004383 (2014).

92. Wang, G., Sarkar, A., Carbonetto, P. & Stephens, M. A simple new approach to variable selection in regression, with application to genetic fine mapping. J. R. Stat. Soc. Series B Stat. Methodol. 82, 1273–1300 (2020).

93. Wallace, C. A more accurate method for colocalisation analysis allowing for multiple causal variants. PLoS Genet. 17, e1009440 (2021).

94. Zheng, J. et al. Phenome-wide Mendelian randomization mapping the influence of the plasma proteome on complex diseases. Nat. Genet. 52, 1122–1131 (2020).

